# Performance of the Heatwave ED Forecast Tool in the Emergency Department Network of Ticino Canton, Switzerland

**DOI:** 10.64898/2026.10.11.26365239

**Authors:** Matteo Paganini, Ilaria Costantini, Ilenia Mascherona, Simone Cosci, Laura Di Domenico, Ana Maria Vicedo-Cabrera, Roberta Petrino

## Abstract

**Objectives:** Heatwaves drive surges in acute care demand, yet heat-health action plans (HHAPs) lack tools to scale emergency care responses. This study validated a forecasting tool for heatwave- related emergency department (ED) demand across four clusters comprising the emergency care network of Ticino Canton, Switzerland.

**Methods:** This retrospective, ecological, time-series study modeled daily mean temperature and ED presentations using a distributed lag non-linear model framework with autoregressive and calendar adjustment components. The model was trained on summers (June–August) 2016-2024 and tested on summer 2025, defining heatwaves as daily mean temperature >25 °C for ≥3 consecutive days.

**Results:** A non-linear temperature–demand association was shown across the clusters, with ED visits increasing at higher temperatures. Model fit varied across clusters (R² 0.389–0.672), and the reference model consistently outperformed naïve baselines. During two heatwaves in 2025, prediction errors remained within acceptable ranges. Baseline demand, heatwave occurrence, and model performance varied substantially within the canton.

**Conclusions:** These findings have implications for the design of HHAPs, suggesting that emergency care preparedness should rely on locally calibrated demand forecast rather than standardized activation protocols.

## INTRODUCTION

Global warming, driven mainly by anthropogenic greenhouse gas emissions, is accelerating climate change and is considered a major health threat of the 21^st^ century (1–3). Europe has warmed faster than any other World Meteorological Organization (WMO) region since the 1980s, with annual mean temperatures rising at about twice the global average. This amplification is even stronger in Western/West-Central Europe, where summer temperatures have risen nearly three times faster than the observed global mean warming (∼2.3 °C in West-Central Europe vs ∼0.8 °C globally, 1980–2022) (4,5).

Global warming is associated with increasing frequency and intensity of extreme weather events (EWEs) (6,7), especially heatwaves (8). Defined as prolonged periods of abnormally high temperatures, heatwaves strongly correlate with increased morbidity and mortality, particularly in vulnerable populations (9), and have substantial repercussions on healthcare systems (10). Heatwaves also generate excess acute care demand and are correlated with increased use of Emergency Medical Services (EMS) and Emergency Departments (EDs), reported in different continents (11–20) as well as in Europe (21–23).

Heat-Health Action Plans (HHAPs) are intended to mitigate the adverse effects of heatwaves on affected populations. Activated by Early Warning Systems, most often linked to meteorological forecasts, HHAPs trigger actions spanning risk communication, protection of vulnerable groups, and preparedness of health and social care services (24,25). In Europe, the implementation of HHAPs was catalyzed by the 2003 heatwave (26); however, the extent to which health services–especially emergency care–are explicitly integrated remains scattered (24,27). This gap is crucial, as heatwave- related patient surges can compromise emergency care delivery, similarly to other EWEs or disasters triggered by natural hazards (12), and may increase in-ED mortality (28). A potential bridge between meteorological EWEs and scalable healthcare actions is health surveillance, yet it currently remains a marginal component of HHAP activation and modulation in Europe, as noted in the WHO’s 2021 HHAPs review (29). Another approach is to develop models that predict emergency care demand when coupled with meteorological forecasts, enabling scalable adaptations and tailored responses to support surge-capacity planning. A previous study developed a predictive model generating ED attendance accounting for the effect of heatwaves using data from the University Hospital of Bern, Switzerland (30). Since Switzerland’s mean temperature has increased by 2.9 °C compared with the pre-industrial period up to 2024 (31), the country is at serious risk of more frequent heatwaves in the near future, especially in warmer regions in the country such as the Ticino canton (31). This study, therefore, aimed to validate the Bern heatwave–ED predictive model in the Ticino Canton, a warmer region with explicit national prioritization for heat-health risk.

## METHODS

### Study design, context, and setting

This retrospective, ecological, time-series study was conducted to quantify the association between ambient temperature (and short-term temperature dynamics) and daily ED presentations, using the “*forecasting tool for hospital demand during heat periods*” previously developed using data from the Inselspital in Bern, Switzerland (30).

Switzerland has a federally organized health system in which compulsory basic health insurance provides nationwide coverage, while the cantons hold primary responsibility for healthcare provision, including hospital planning and oversight. In Ticino Canton, the southernmost canton and bordering Italy, the public acute-care hospital network is coordinated through the Ticino Canton Hospital Trust (Ente Ospedaliero Cantonale – EOC), a cantonal public institution that manages and integrates the main public hospitals serving the resident population and cross-border catchment. Emergency care is delivered through an integrated prehospital–hospital pathway: prehospital responses are activated via the national emergency number, stabilized, and transferred where appropriate. Within the EOC network, 24/7 ED capability is concentrated in four districts (Lugano, Bellinzona, Locarno, and Mendrisio), complemented by proximity first-aid posts in peripheral valleys to support access and safe onward referral. Analyses were therefore performed separately in the four districts constituting the cantonal emergency care network.

### Data sources

#### ED demand data

Four ED clusters were created to allow pairing with meteorological data in case of areas with a mix of emergency care structures, mirroring the healthcare network districts, as follows:

- Bellinzona: including the "San Giovanni" Hospital in Bellinzona, and first aid posts in Acquarossa and Faido;
- Lugano: including the Lugano Regional Hospital and the “Ospedale Italiano” of Lugano (a facility incorporated with the Regional Hospital in 2024);
- Mendrisio: including the "Beata Vergine" Hospital in Mendrisio;
- Locarno: including the “La Carità” Hospital in Locarno

For each cluster, the daily aggregated number of ED presentations was derived from routinely collected hospital data. The available time coverage spanned September 2015 to August 2025; the analytic dataset was restricted a priori to 01 January 2016 through 31 August 2025 to ensure consistent year coverage and complete lagged predictors.

#### Meteorological data

Daily meteorological observations were retrieved from MeteoSwiss open-data web sourced database (32), using station-level time series. ED clusters were linked to pre-specified meteorological stations (monitoring towers) to create an ED cluster-specific exposure series, as follows: Bellinzona to Grono station; Lugano to Lugano station; Mendrisio to Stabio station; and Locarno to Locarno Monti station.

### Exposure definition

The primary exposure was daily mean ambient temperature (T_mean), modeled as a non- linear, lagged exposure using a distributed lag non-linear model (DLNM) framework over lags 0–3 days. For targeted forecasting evaluation during hot waves, we used the operational definition of MeteoSwiss, identifying a heatwave as T_mean > 25 °C for at least 3 consecutive days. A descriptive analysis was conducted for the summer period (1 June–31 August) from 2016 to 2025. For each year, the total number of ED visits was calculated. Continuous variables were summarized as mean and standard deviation (SD) or median and interquartile range (IQR), as appropriate according to their distribution. Heatwave periods during the test year (2025) were identified through graphical inspection of the temperature time series.

### Statistical modeling

#### Core regression framework

We used the tool’s time-series regression framework with an autoregressive component to model daily ED presentations in each cluster. The model was fitted assuming Gaussian errors, consistent with the original Bern forecasting model. For each cluster, the outcome was the daily ED visit count, with the following predictor blocks:

- Autoregressive demand component: ED visit counts in the previous 7 days (lags 1–7) to account for short-term persistence in demand;
- Calendar adjustment: indicator variables for day of week and public holidays;
- Pandemic-period adjustment: an indicator capturing pandemic years (2020-2022) to account for the structural disruption in ED use during COVID-19;
- Temperature component (non-linear and lagged): the association between T_mean and ED visits was modeled as non-linear and distributed over short lags (see below).

#### Temperature–lag specification (distributed lag non-linear model)

Temperature was modeled using natural cubic splines and a short-distributed lag structure. We implemented the tool’s default parameterization: a spline with internal knots at the 10th, 50th, and 90th percentiles of the temperature distribution (corresponding to 4 degrees of freedom for the exposure–response curve), and a lag window of 3 days (evaluating effects over lag 0 to lag 3). This specification yields both an exposure–lag–response surface and a cumulative temperature effect across lags.

### Model development and evaluation

Following the tool’s intended use, model development and evaluation were restricted to the summer period (01 June–31 August) of each year. Therefore, a temporal hold-out strategy aligned with real-world deployment was adopted:

- Training period: summers (01 June–31 August) from 2016–2024;
- Test period: summer 2025 (01 June–31 August).

Model performance was evaluated on the held-out test summer by comparing predicted and observed daily counts in each ED cluster, and by examining performance specifically during heatwave periods as defined above (T_mean > 25 °C for ≥3 consecutive days). In particular, we computed two error metrics: Root Mean Squared Error (RMSE) and Mean Absolute Error (MAE).

### Reporting of estimates

For each ED site, the following estimates were reported: (i) the estimated non-linear association between temperature and ED cluster visits at individual lags, (ii) the cumulative temperature–demand association across the lag window, and (iii) predictive performance in the test period, including heatwave-focused assessment.

### Data governance and ethics aspects

All analyses were conducted on aggregated daily counts and meteorological time series. Data were handled in accordance with applicable institutional and data-protection requirements. The study was exempted from formal review by the local ethics committee (CE TI 4951–Req-2025-01236).

## RESULTS

### Descriptive overview

Across the decade, the ED demand in the Ticino Canton showed an overall increasing trend, with a clear transient reduction during 2020–2022 (pandemic years). In the training period (summers 2016–2024), mean (SD) daily ED clusters presentations were 89.5 (18.7) in Bellinzona, 64.0 (13.8) in Locarno, 113.0 (19.5) in Lugano, and 52.2 (10.2) in Mendrisio. In the test period (summer of 2025), corresponding values were 108.0 (13.0), 72.9 (10.7), 95.8 (10.2), and 56.2 (8.8), respectively. An increasing trend can be noted in 2025, except in Lugano (Figure 1; Table 1).

**Figure 1.**
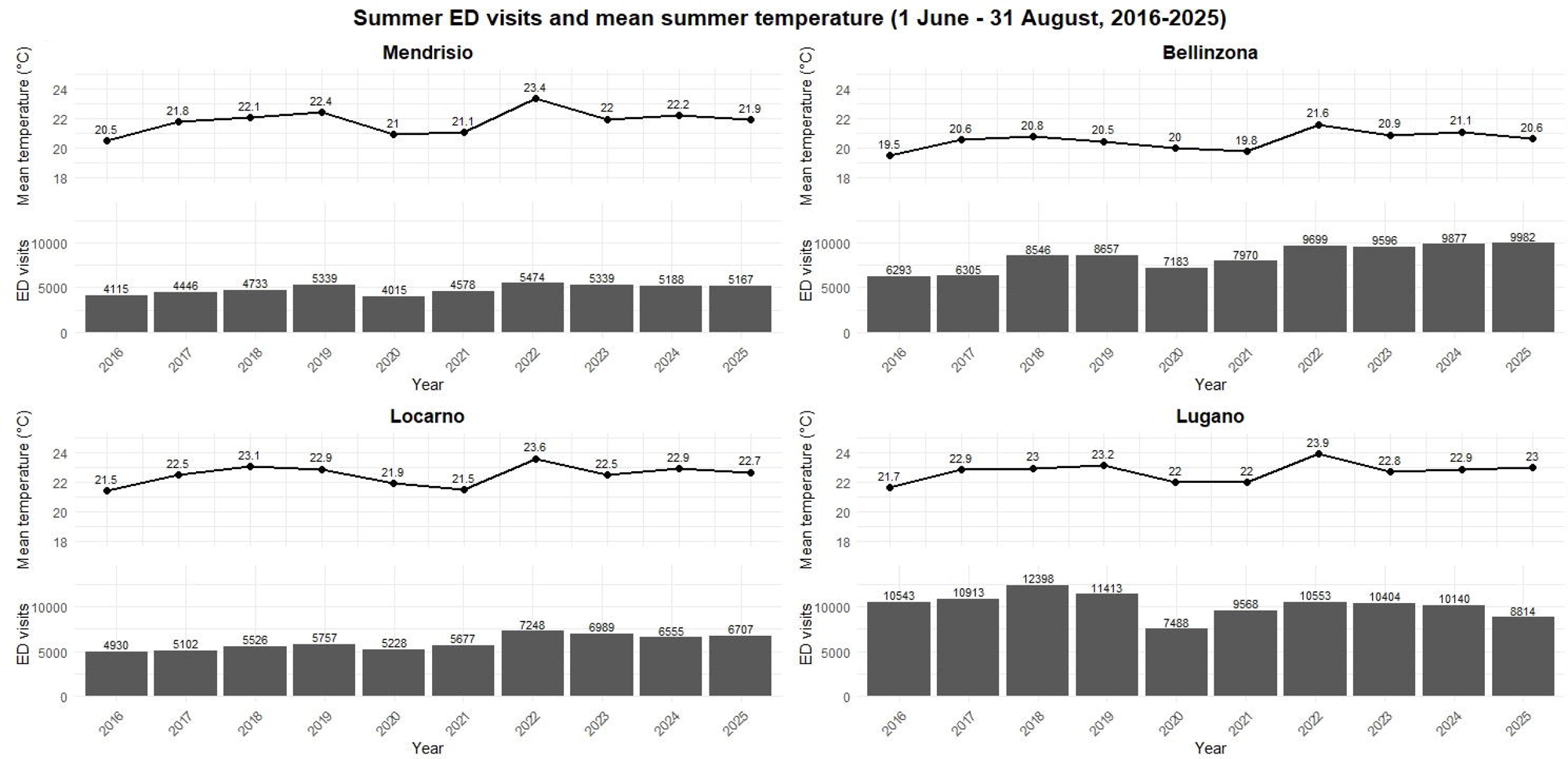
Total number of Emergency Department (ED) visits and mean temperature during the summer period (1 June–31 August) from 2016 to 2025 in the four ED clusters (Mendrisio, Bellinzona, Locarno, and Lugano) of Ticino Canton, Switzerland. Bars represent the total number of ED Visits recorded each summer, with values displayed above each bar. A marked reduction in ED Visits is visible in 2020-2021, likely reflecting the impact of the COVID-19 pandemic, followed by a gradual increase in subsequent years in most locations. On the above continuous black line, average temperature registered during each year’s summer are reported. These data provide the temporal context for the forecasting models developed in this study. Ticino Canton, Switzerland, 2016-2025.

**Table 1.**
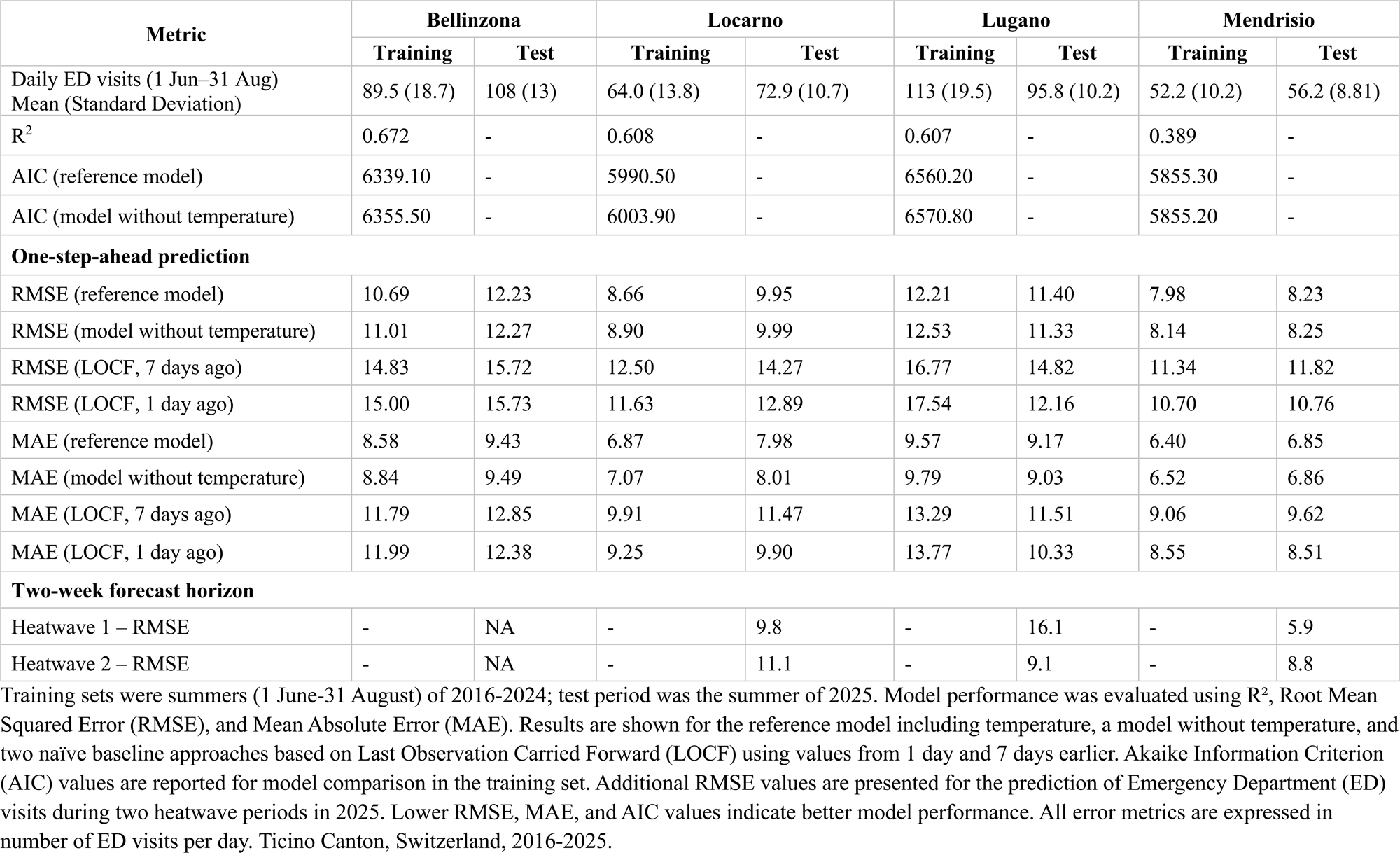
Forecasting performance across clusters in training and test periods across four Emergency Department clusters.

### Temperature–demand association

Across clusters, daily mean temperature showed a non-linear association with ED presentations, with heterogeneous temperature–demand patterns. ED demand increased at higher temperatures along with uncertainty (Figure 2). Lag-specific functions are reported in the Supplementary File 1 for model transparency.

**Figure 2.**
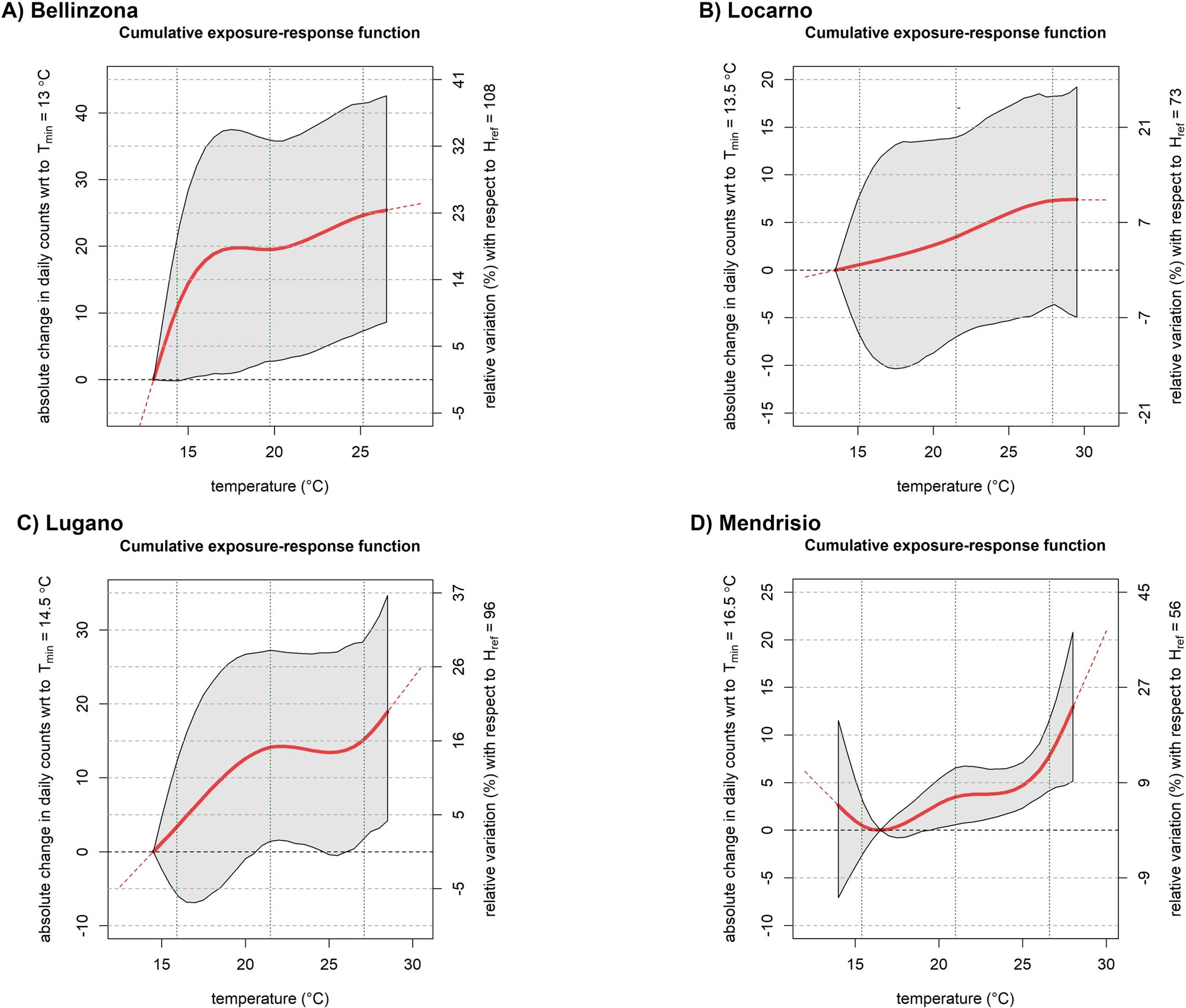
Exposure-response function estimated for the four Emergency Department clusters. (A) - Bellinzona; (B) - Locarno; (C) - Lugano; (D) - Mendrisio. For each cluster, the panel shows the cumulative function, expressed as the absolute change in daily hospital visits according to temperature, with the red line indicating the estimated association and the shaded area the 95% confidence interval; the right y-axis reports the corresponding relative variation (%) with respect to the reference level. Ticino Canton, Switzerland, 2016-2025.

### Heatwave periods in summer 2025

Using the operational definition of heatwave (daily mean temperature >25 °C for ≥3 consecutive days), two heatwave periods were identified in summer 2025 in three clusters, with small date shifts reflecting local meteorological conditions. In Locarno, heatwaves occurred on 26–29 June 2025 and 9–17 August 2025. In Lugano, heatwaves occurred on 24–30 June 2025 and 9–15 August 2025. In Mendrisio, heatwaves occurred on 28–30 June 2025 and 10–14 August 2025. No heatwave periods were identified in Bellinzona during summer 2025. For more details, see Supplementary File 1.

### Model fit and forecasting performance

Model fit, assessed on the training data, showed substantial explanatory capacity in Bellinzona (R²=0.672), Locarno (R²=0.608), and Lugano (R²=0.607), whereas a lower proportion of variance was explained in Mendrisio (R²=0.389). In Bellinzona, Locarno, and Lugano clusters the temperature- augmented model provided a better fit than the model without temperature, as reflected by lower AIC values (Bellinzona: 6339.1 vs 6355.5; Locarno: 5990.5 vs 6003.9; Lugano: 6560.2 vs 6570.8). In Mendrisio, model fit was similar with and without temperature (AIC 5855.3 vs 5855.2) (Table 1).

In the test summer 2025, one-step-ahead prediction errors were stable across clusters. Test-set RMSEs for the reference (temperature-augmented) model were 12.23 (Bellinzona), 9.95 (Locarno), 11.40 (Lugano), and 8.23 (Mendrisio). Corresponding MAEs were 9.43, 7.98, 9.17, and 6.85.

Compared with a model excluding temperature, differences in overall test-set error were small (RMSE: 12.27, 9.99, 11.33, and 8.25, respectively), indicating that the incremental contribution of temperature to overall summer forecasting accuracy was modest (Table 1).

Across clusters, the reference model outperformed naïve last-observation-carried-forward (LOCF) baselines based on ED demand alone. In the test set, RMSEs for the 1-day LOCF ranged from 10.76 to 15.73 and for the 7-day LOCF from 11.82 to 15.72, consistently higher than the reference model RMSEs (Table 1).

### Forecast errors during heatwaves

During the identified heatwave periods in 2025, prediction errors (evaluated on a two-week horizon) remained within the same order of magnitude as overall summer performance, although variability differed by cluster and event. For Locarno, RMSE was 9.8 during the late-June heatwave and 11.1 during the August heatwave. For Lugano, RMSE was 16.1 during the late-June heatwave and 9.1 during the August heatwave (a representative example is available as Figure 3). For Mendrisio, RMSE was 5.9 during the late-June heatwave and 8.8 during the August heatwave (Table 1). All the forecasts generated during the different heatwaves are detailed in the Supplementary File 1.

**Figure 3.**
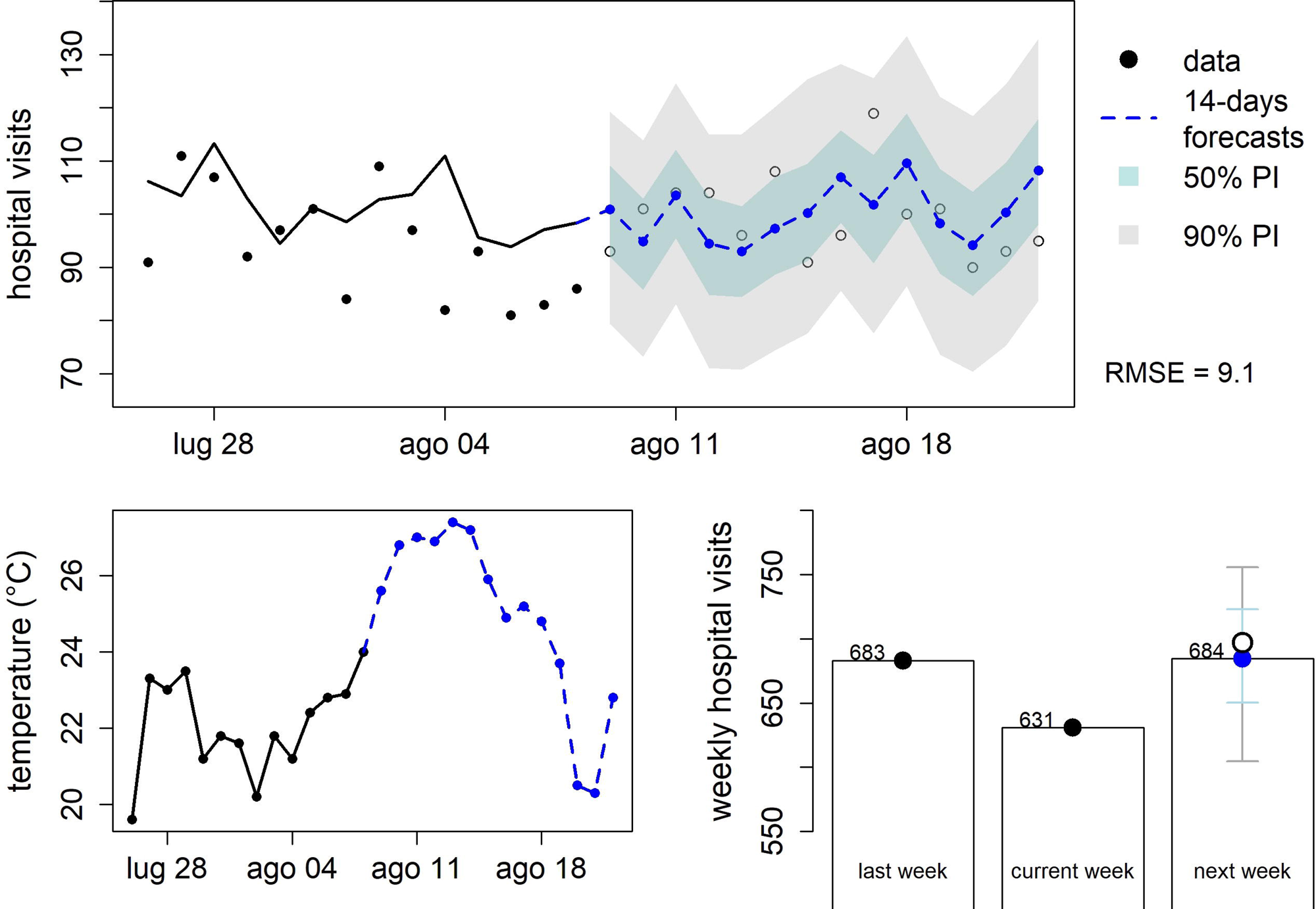
**Forecast example: Forecast of emergency department visits in Lugano during the August 2025 heatwave (HW2)**. The upper panel shows observed daily hospital visits from 28 July to 8 August 2025 (black points and line) and 14-day forecasts from 9 to 22 August 2025 (blue dashed line) with 50% and 90% prediction intervals. The forecast period includes the heatwave observed in Lugano from 9 to 15 August 2025. The lower left panel shows the corresponding daily mean temperature (°C). The lower right panel summarizes weekly hospital visits, comparing the previous week, the current week, and the forecast for the following week. Ticino Canton, Switzerland, 2016-2025.

We compared forecast errors of a model with or without temperature in a cross-validation analysis, training the two model versions on all summers except for one year used as test set (Supplementary File 1). We focused on high-temperature periods, considering forecast windows with a duration of three weeks and an average temperature above 25 °C. For a one-week forecast horizon, the two models showed similar median RMSE (in all clusters, except for Bellinzona, where the number of selected time windows was limited). However, at longer forecast horizons (two to three weeks ahead), the model including temperature exhibited lower errors than the model excluding temperature. Although the improvements in forecasting performance when incorporating temperature were modest, they were observed consistently across clusters.

## DISCUSSION

This study validates a tool for forecasting heatwave-related daily ED presentations across the four ED networks of Ticino Canton in Switzerland and provides implementation-relevant insights for integrating emergency care initiatives into HHAPs. Ticino Canton was selected for the validation because of its climate-risk profile and its designation as a high-priority region for climate adaptation by Switzerland’s environmental authority (33). Switzerland’s mean temperature has increased by approximately 2.9°C relative to the pre-industrial period up to 2024 (31), and ED presentations in Ticino rose by 8.4% during the 2015 heatwave (34). Under a worst-case warming scenario of roughly +4°C (RCP 8.5) (35), heatwaves are expected to become more frequent and intense, with a corresponding increase in acute-care demand. The cluster-based analysis allowed us to estimate the exposure-response function for each of the four ED networks, linking temperature with ED visits.

Although the shape and magnitude of the association varied across clusters, the results consistently showed greater increases in ED visits corresponding to particularly high temperature values (Figure 2). These findings align with those from the Bern case study (30) and suggest a correlation between heatwaves and increased ED demand.

As a key safety net in modern societies, emergency medicine works to ensure that no one is left behind while confronting overcrowding. This persistent global problem is a sensitive indicator of health-systems strain (36) and reflects the combined effects of input burden, internal throughput, and output constraints (37). Heatwaves often increase presentations (input), so even moderate surges can have disproportionate operational impacts and downstream harms when throughput and output are already near capacity (due to crowding). This aligns with broader evidence that ED crowding strongly influences system performance, as demand variability interacts with structural and procedural bottlenecks to produce episodic overload (37). In this context, the long-term rise in ED demand observed in the canton across the years supports treating input as variable and highlights the need for tools that can forecast fluctuations in emergency care demand to inform surge capability decisions.

This work adopted a cluster-based modeling strategy, assuming that ED demand, heat exposure, and local operational context may vary even within a single canton. In Ticino, such variability was evident: baseline summer ED presentations differed substantially across the four clusters, potentially reflecting differences in population age and morbidity, and care-seeking behavior. Heatwave occurrence during the test summer of 2025 was also uneven, with two heatwave periods in Locarno, Lugano, and Mendrisio, respectively, but none in Bellinzona. This within-canton heterogeneity likely reflects microclimatic and local exposure differences captured by station-based temperature series. Such differences can further amplify demand variability through variation in local heat effects and, consequently, in heat-health and heat-healthcare systems interactions, as reported in multiple settings (9,14,16,38). Heterogeneity was also evident from model performance, with similar R² in Bellinzona, Locarno, and Lugano (0.607–0.672) but notably lower in Mendrisio (0.389) (Table 1). In particular, the lower R^2^ in Mendrisio should not be interpreted solely as model failure, but may reflect a combination of lower statistical power due to smaller daily volumes, unmeasured local drivers of demand, or contextual factors. For example, Mendrisio is the southernmost part of the canton, has a smaller hospital, and lies close to the Italian border, all of which could influence presentation volumes. A “single-canton, single-model, single-planning” approach therefore risks averaging away locally meaningful differences and may mislead executives and operational users. Cluster-specific tools with strong customization capabilities may better match how emergency care networks plan resources and respond to warnings under local constraints and population needs.

Another aspect that warrants critical evaluation is the usefulness of temperature for both model performance and interpretation. Temperature contributed little to model accuracy, as RMSE differences between the temperature and no-temperature variants were close to negligible (Table 1). Adding temperature improved model fit in Bellinzona, Locarno, and Lugano (lower AIC) but not in Mendrisio. The reference model also achieved similar error ranges across clusters and consistently outperformed naïve LOCF baselines. Two factors may explain these patterns: (i) mean summer error metrics are dominated by non-extreme days, so any benefit from temperature may appear mainly in the upper tail of heat exposure and be diluted in average measures; (ii) autoregressive and calendar components may already capture much of the predictable variation, leaving limited room for improvement from a single environmental covariate. Although the DLNM framework estimated a short-lag effect (Supplementary File 1) consistent with trends reported in the literature on acute and delayed clinical effects of heatwaves (9,39–45), these results are unstable and should be interpreted with caution, as potentially due to model artifacts. Overall, temperature may add more to interpretability than to season-wide average accuracy by helping anticipate high-impact days and support scalable decisions. In Ticino hospitals, where no routine ED demand forecasting is currently in use, either model variant (with or without temperature) may provide more informative and actionable guidance for activating patient-surge plans than simple LOCF approaches.

From a public health perspective, this work supports a better integration of emergency care into HHAPs, which to date has been implemented inconsistently across Europe (24). Recent works highlight fragmentation of preparedness measures across sectors (46) and gaps in actionable triggers for EMS and ED operations (20,47–49). Although specific analyses of implementation barriers remain limited, the tool validated in this study offers a pragmatic bridge between meteorological warnings and scalable actions in emergency care. Operationally, meteorological forecasts processed through the model could inform escalation decisions in the days immediately before a heatwave and support flexible planning for subsequent days. Actions could be organized using an emergency care operations flow model (*input-throughput-output*) (50) or surge science principles (*staff*, *stuff*, *structure* and *system*–the disaster medicine’s “4-S approach”) (51), as suggested by seminal literature (10,20) or qualitative research (47–49). The cluster-specific design of the tool also complements and extends prior work forecasting heat-related demand from a single urban Swiss ED setting (30) and provides evidence of within-canton heterogeneity that is likely relevant in other contexts. In fact, a single national or region-wide forecast tool may be easier to develop and implement, but it is unlikely to be appropriate. Instead, a national or canton-wide plan could coordinate structured multiagency cooperation, while preserving granular forecast tools and responses, avoiding overly generic–and potentially resource-wasting–activations across the canton. Finally, local calibration could further improve model fit in each cluster and better capture local demand responses. The model can be retrained after each summer, allowing the exposure-response function to be refined as the climate evolves. The tool could also be embedded in a learning system, with event-specific deviations triggering investigation of unmeasured drivers (e.g., holiday mobility, cross-border care-seeking, air pollution, capacity constraints) to guide iterative model refinement.

### Limitations

Several limitations should be considered when interpreting and applying the model. First, the outcome is all-cause, aggregated daily ED presentations. Without patient-level data (such as diagnosis or acuity) more granular modeling and mechanistic interpretation are limited, and operational users should not assume that predicted increases reflect a specific clinical syndrome. Second, exposure was defined to mirror the indicators (mean daily temperature) and heatwave definition used by the national meteorological authority (MeteoSwiss), using one station per cluster. Humidity, night-time minima, thermal indices (e.g., UTCI/WBGT/Humidex), and air quality were not included, although these factors may modify heat-related health impacts and could improve event-level performance. Third, the weaker fit and minimal AIC gain from temperature in Mendrisio suggest that local demand may be influenced by drivers not captured in the current specification. Future models could refine these aspects by incorporating additional, locally weighted predictors and calibrating the model accordingly. Fourth, the analysis focused on the summer season; shoulder seasons and compound extremes were not assessed. Finally, the evaluation used observed meteorological data. Because operational deployment will rely on forecast temperatures, prospective validation using forecast inputs and real- time calibration is an essential next step.

## Conclusions

This work validated a tool to forecast heatwave-related ED demand in Ticino Canton, Switzerland. The model suggests an association between heatwaves and higher ED presentations, with heterogeneous patterns across clusters, supporting cluster-specific calibration before implementation. Operationally, the tool could support integrating emergency care initiatives into HHAPs to preserve resilience during heatwaves, while enabling local customization, calibration, and scalable responses.

## Data Availability

All data produced in the present study are available upon reasonable request to the authors

## Notes

### Competing Interest Statement

The authors have declared no competing interest.

### Author Declarations

Ethics committee of Ente Ospedaliero Cantonale (EOC) waived ethical approval for this work (CE TI 4951 Req-2025-01236).

