## Supplementary Material 1 for "Performance of the Heatwave ED Forecast Tool in the Emergency Department Network of Ticino Canton, Switzerland"

##### BELLINZONA (GRONO)

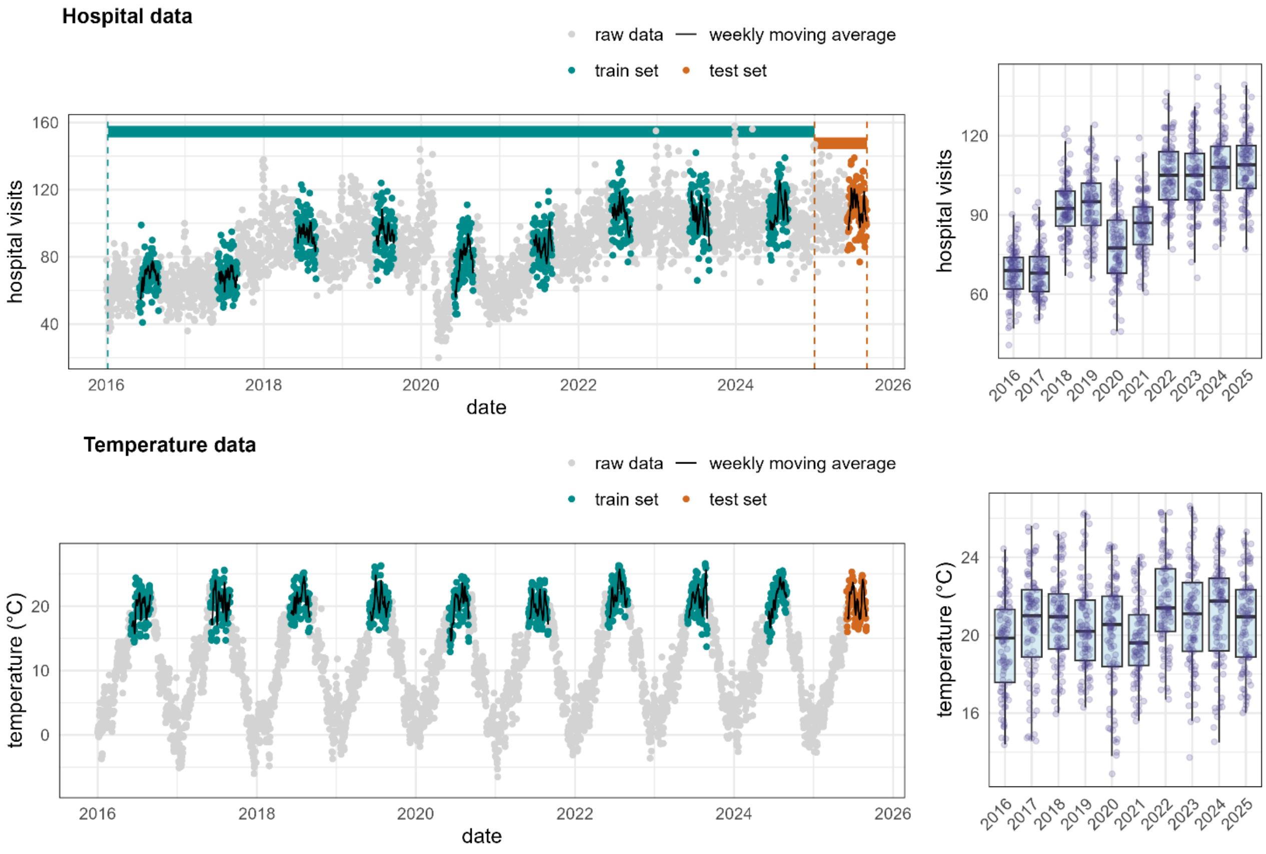


Daily mean temperature (°C) in Bellinzona (Grono station) during the summer period (1 June–31 August) from 2016 to 2025. The dashed line represents the 25 °C threshold; red points indicate days when the temperature exceeded this value, identifying potential heatwave conditions.

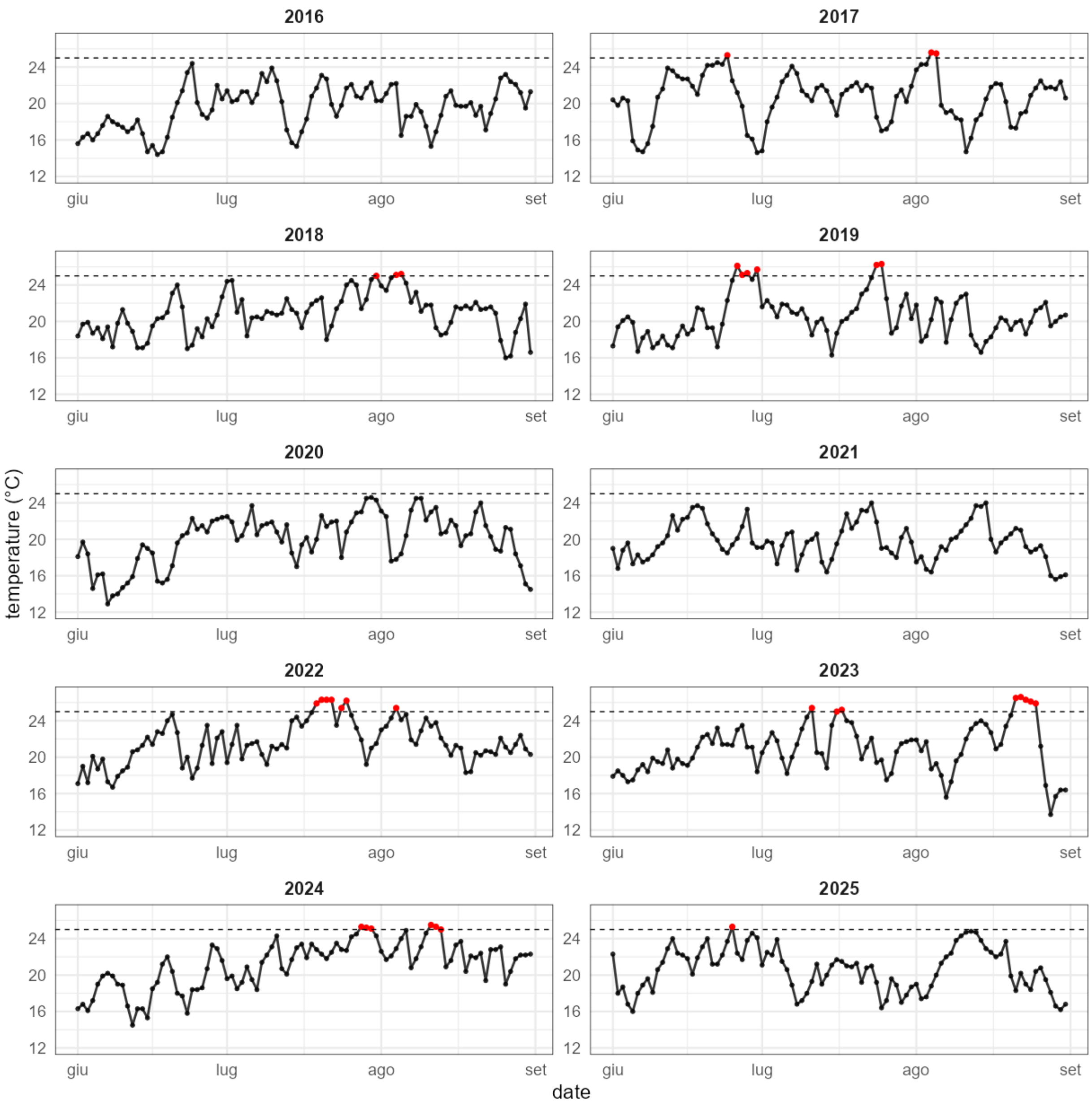


Daily mean temperature (°C) in Bellinzona (Grono station) during the summer period (1 June–31 August) from 2016 to 2025. The dashed line represents the 25 °C threshold; red points indicate days when the temperature exceeded this value.

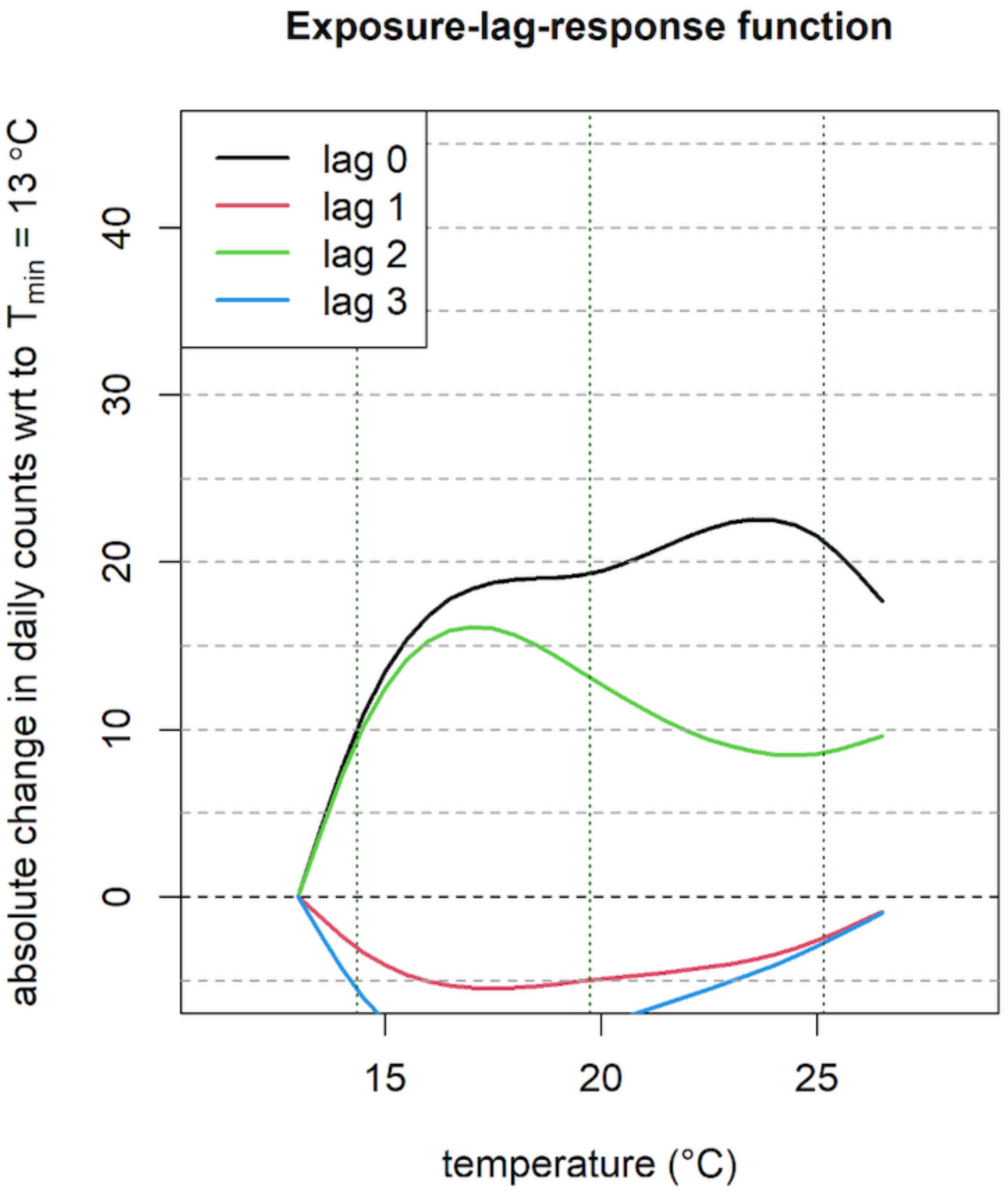


Exposure–lag–response functions describing the association between daily mean temperature and the absolute change in daily emergency department (ED) visit counts in Bellinzona.

*Forecast errors (cross validation). Temperature threshold: 25°C*

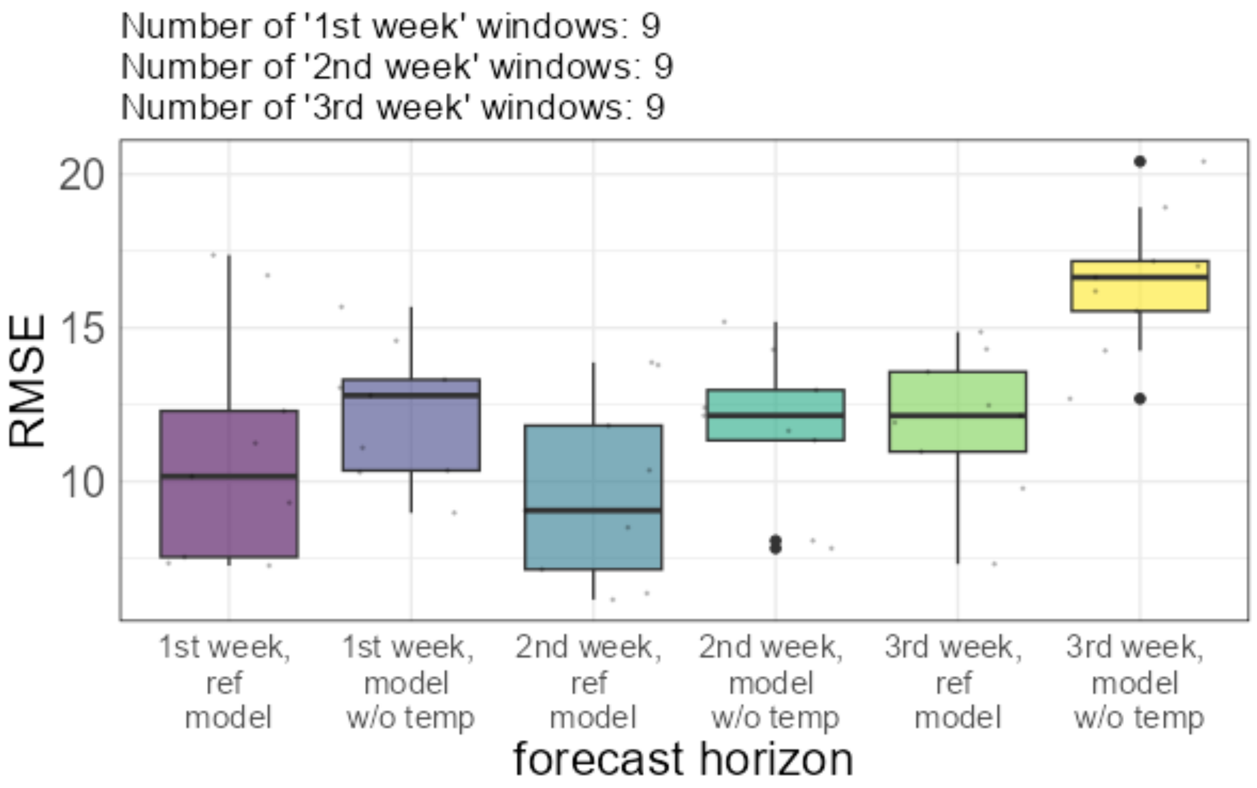


**Comparison of forecast errors for a model with or without temperature.** For each year, we select time-windows of a duration of three weeks, where the average temperature is above a threshold of 25°C. For each year, we generate forecasts for the selected time-windows, using a model trained on all data excluded the year3333 of the test set (cross-validation). We evaluate forecasts errors over a period of 7 days, in the first, second or third week from the starting date of the time-window. We compare the forecast errors of the complete model with a model neglecting temperature as a covariate.

#### LOCARNO (LOCARNO-MONTI)

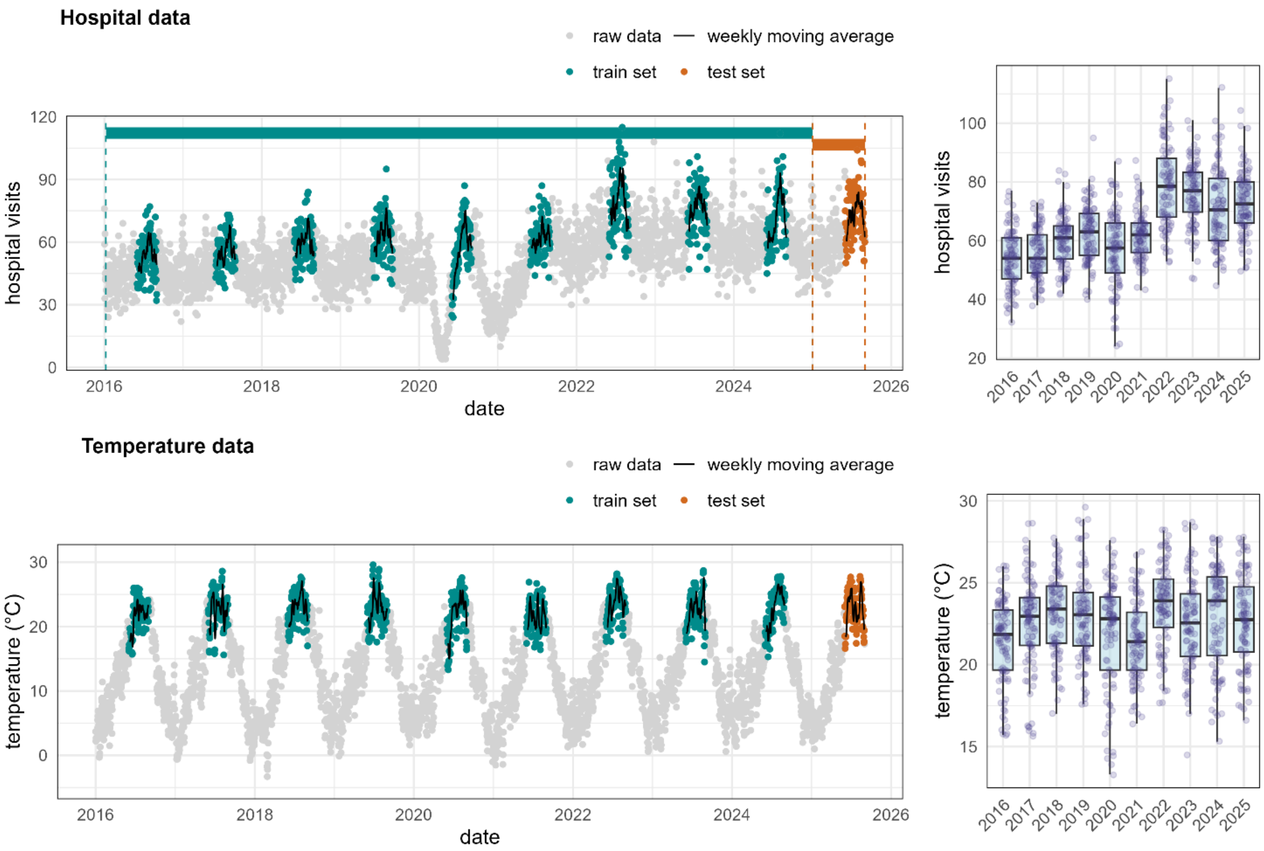


Daily mean temperature (°C) in Locarno (Locarno-Monti station) during the summer period (1 June–31 August) from 2016 to 2025. The dashed line represents the 25 °C threshold; red points indicate days when the temperature exceeded this value, identifying potential heatwave conditions.

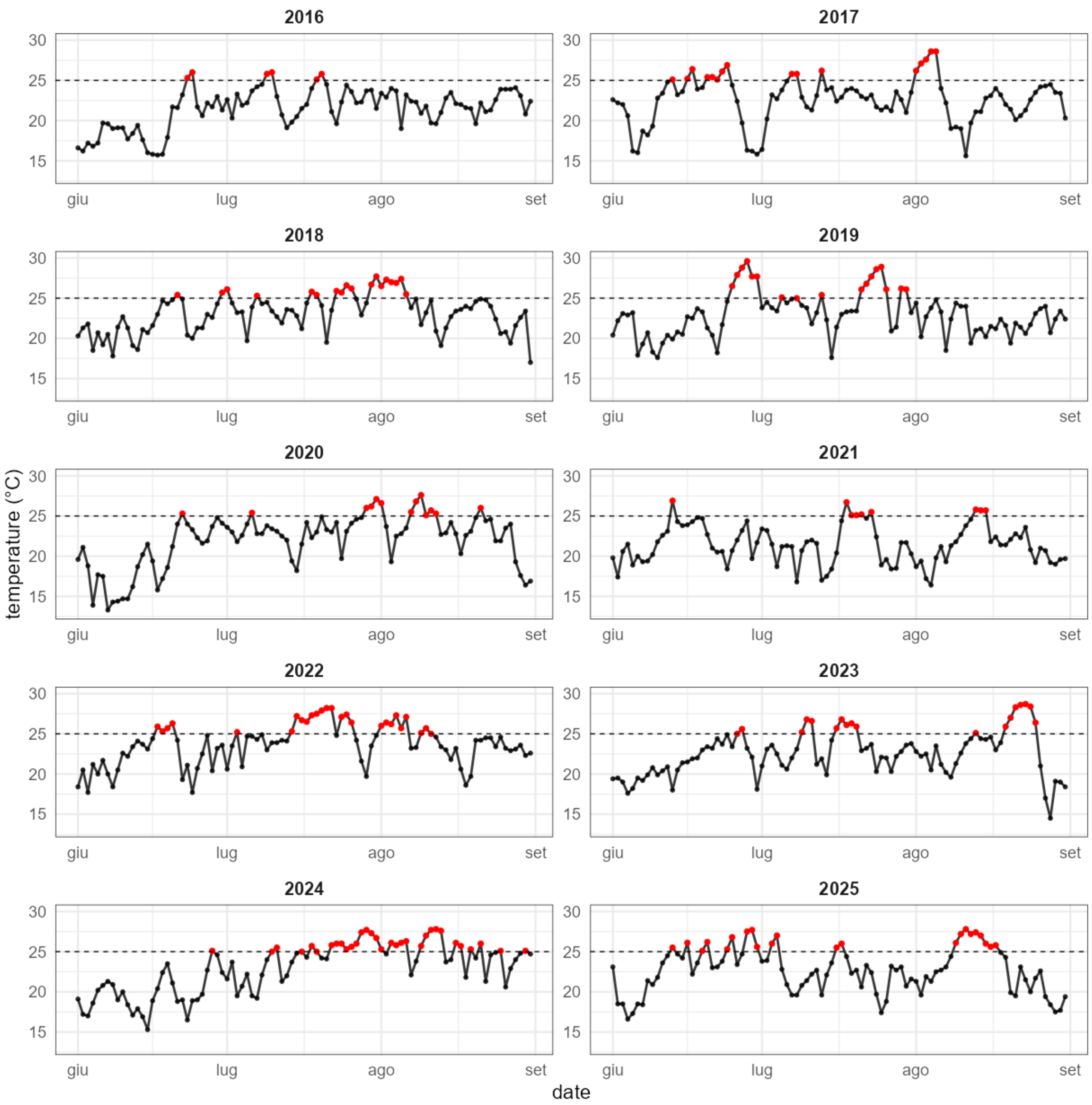


Daily mean temperature (°C) in Locarno (Locarno-Monti station) during the summer period (1 June–31 August) from 2016 to 2025. The dashed line represents the 25 °C threshold; red points indicate days when the temperature exceeded this value.

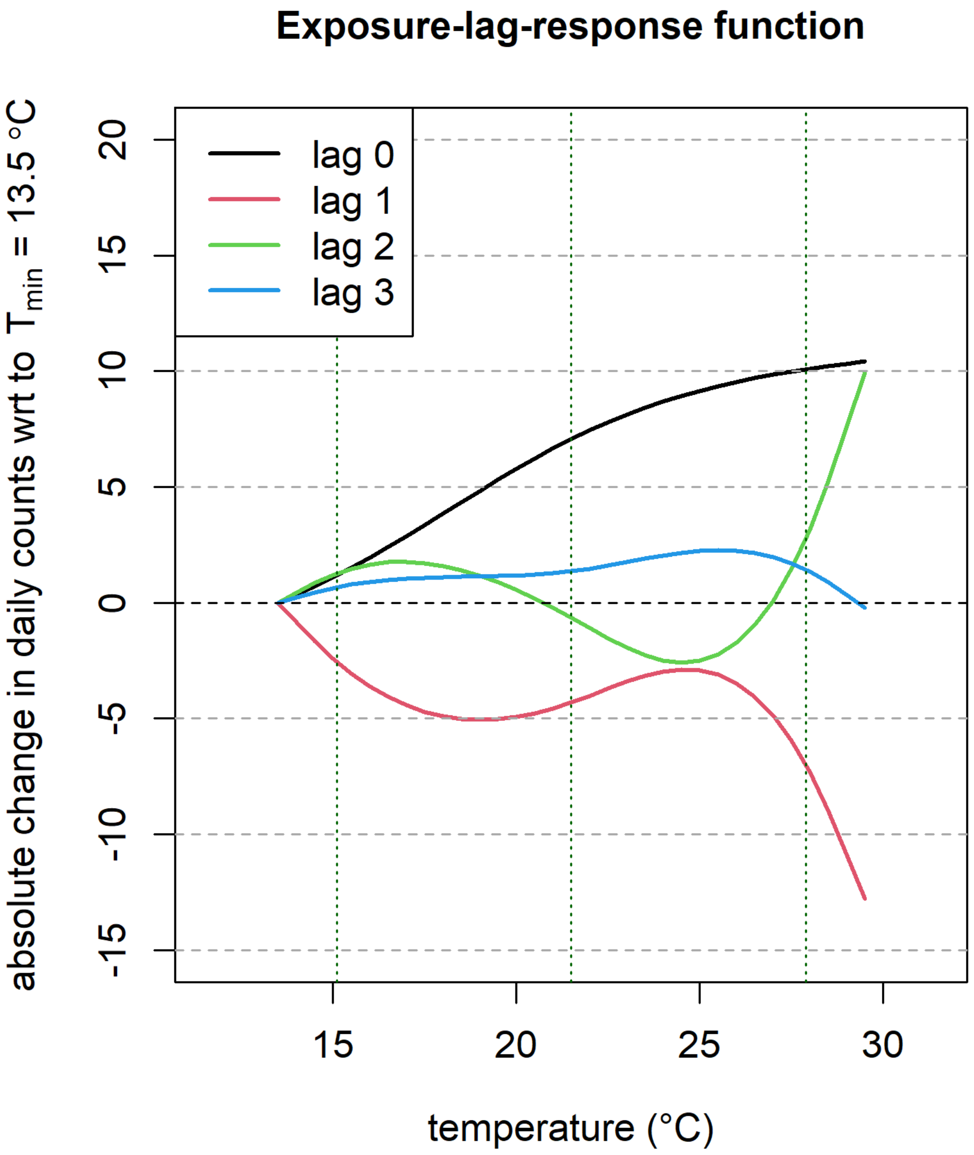


Exposure–lag–response functions describing the association between daily mean temperature and the absolute change in daily emergency department (ED) visit counts in Locarno.

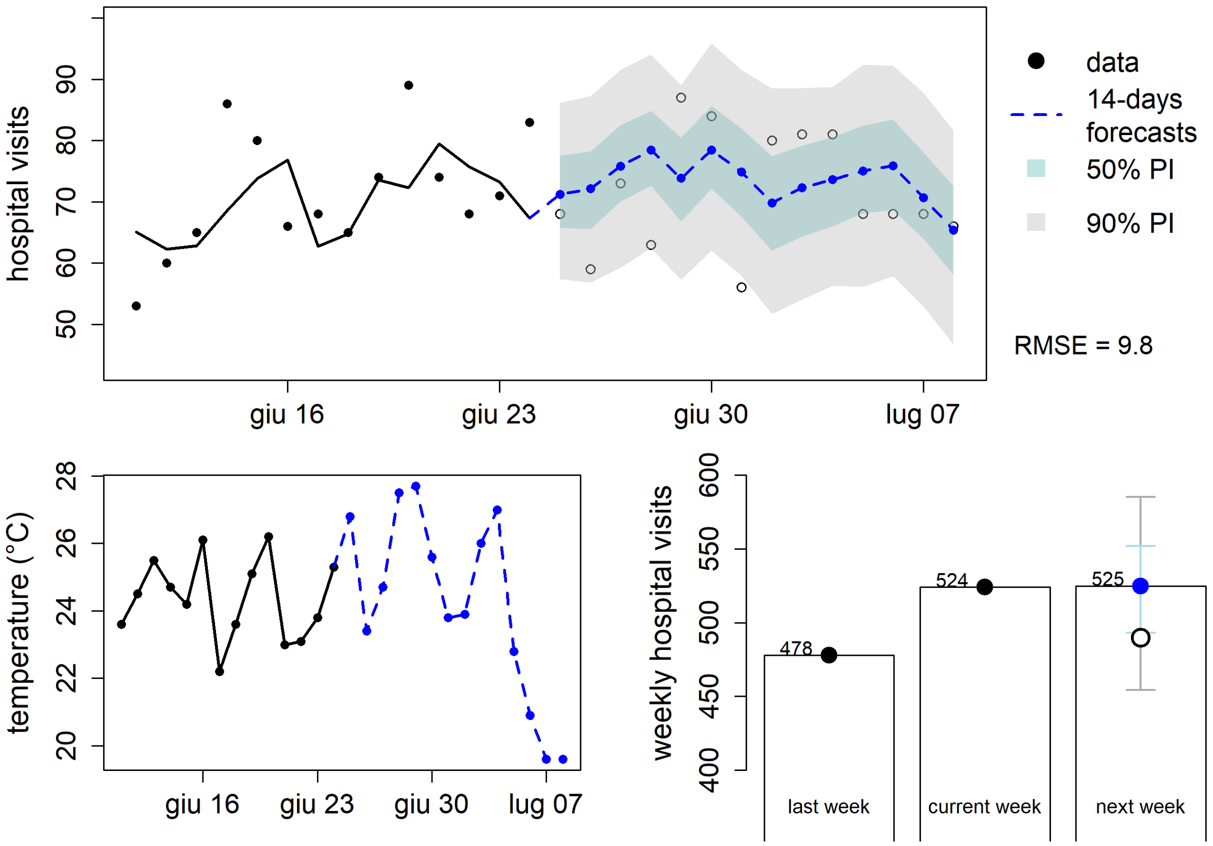


**Forecast of emergency department visits in Locarno during the June 2025 heatwave (HW1).** The upper panel shows observed daily hospital visits from 12 to 25 June 2025 (black points and line) and 14-day forecasts from 26 June to 9 July 2025 (blue dashed line) with 50% and 90% prediction intervals. The forecast period includes the heatwave observed in Locarno from 26 to 29 June 2025. The lower left panel shows the corresponding daily mean temperature (°C). The lower right panel summarizes weekly hospital visits, comparing the previous week, the current week, and the forecast for the following week.

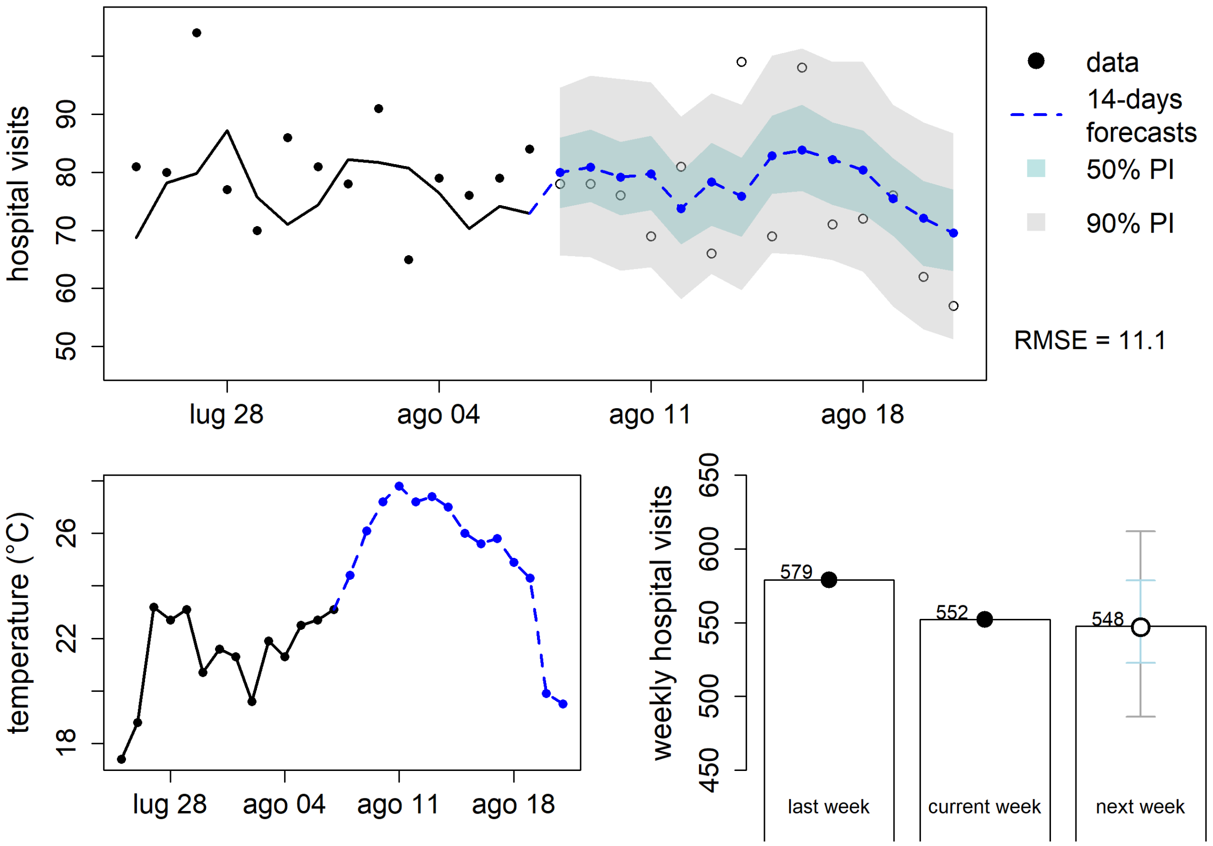


**Forecast of emergency department visits in Locarno during the August 2025 heatwave (HW2).** The upper panel shows observed daily hospital visits from 27 July to 8 August 2025 (black points and line) and 14-day forecasts from 9 to 22 August 2025 (blue dashed line) with 50% and 90% prediction intervals. The forecast period includes the heatwave observed in Locarno from 9 to 17 August 2025. The lower left panel shows the corresponding daily mean temperature (°C). The lower right panel summarizes weekly hospital visits, comparing the previous week, the current week, and the forecast for the following week.

*Forecast errors (cross validation). Temperature threshold: 25°C*

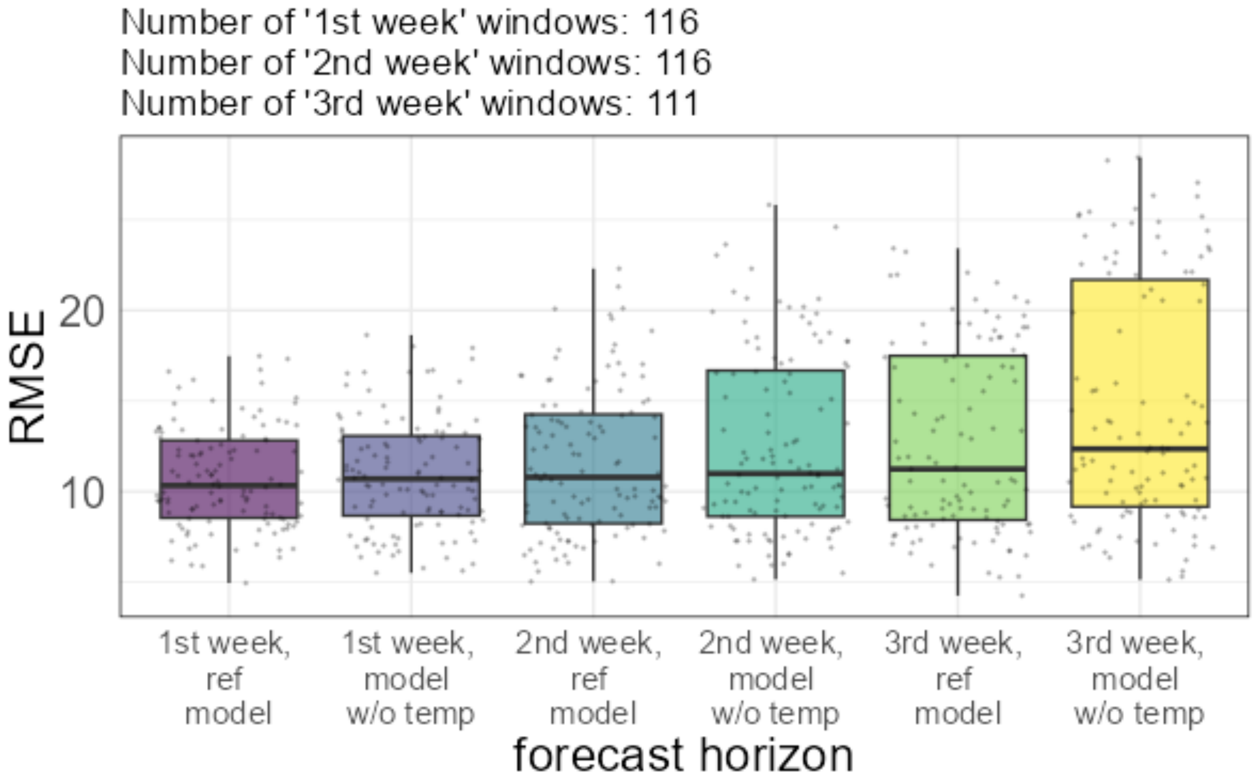

### LUGANO (LUGANO)

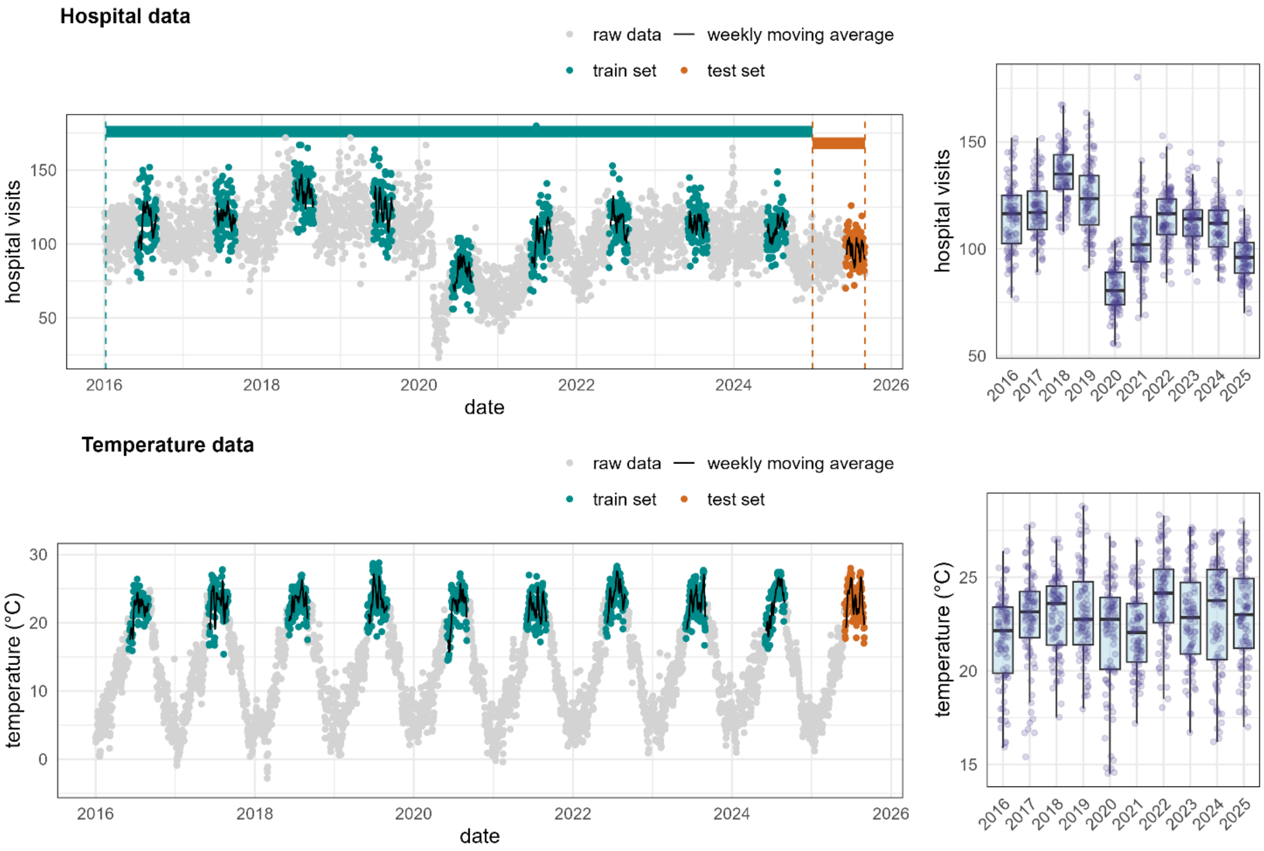


Daily mean temperature (°C) in Lugano (Lugano station) during the summer period (1 June–31 August) from 2016 to 2025. The dashed line represents the 25 °C threshold; red points indicate days when the temperature exceeded this value, identifying potential heatwave conditions.

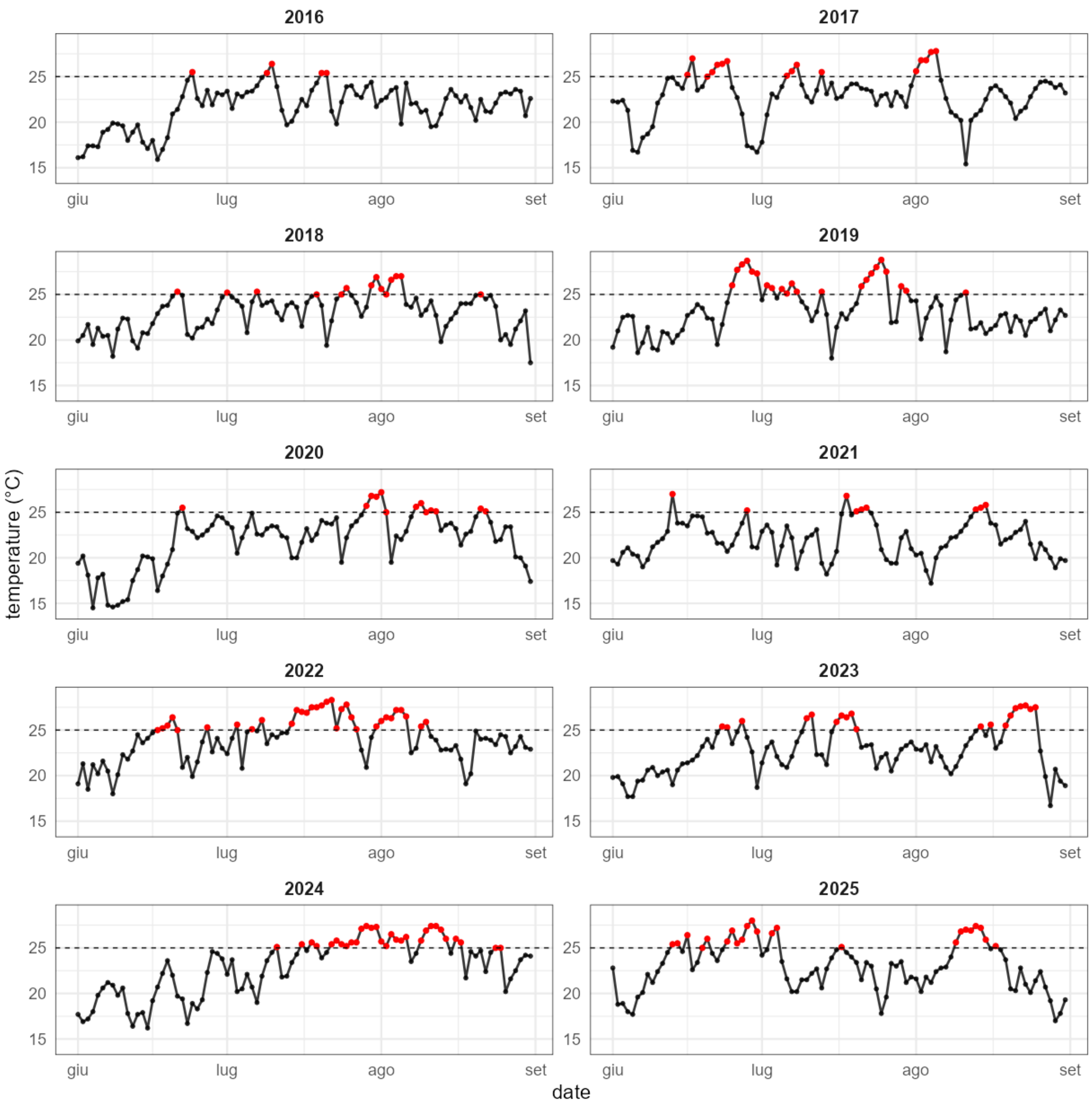


Daily mean temperature (°C) in Lugano (Lugano station) during the summer period (1 June–31 August) from 2016 to 2025. The dashed line represents the 25 °C threshold; red points indicate days when the temperature exceeded this value.

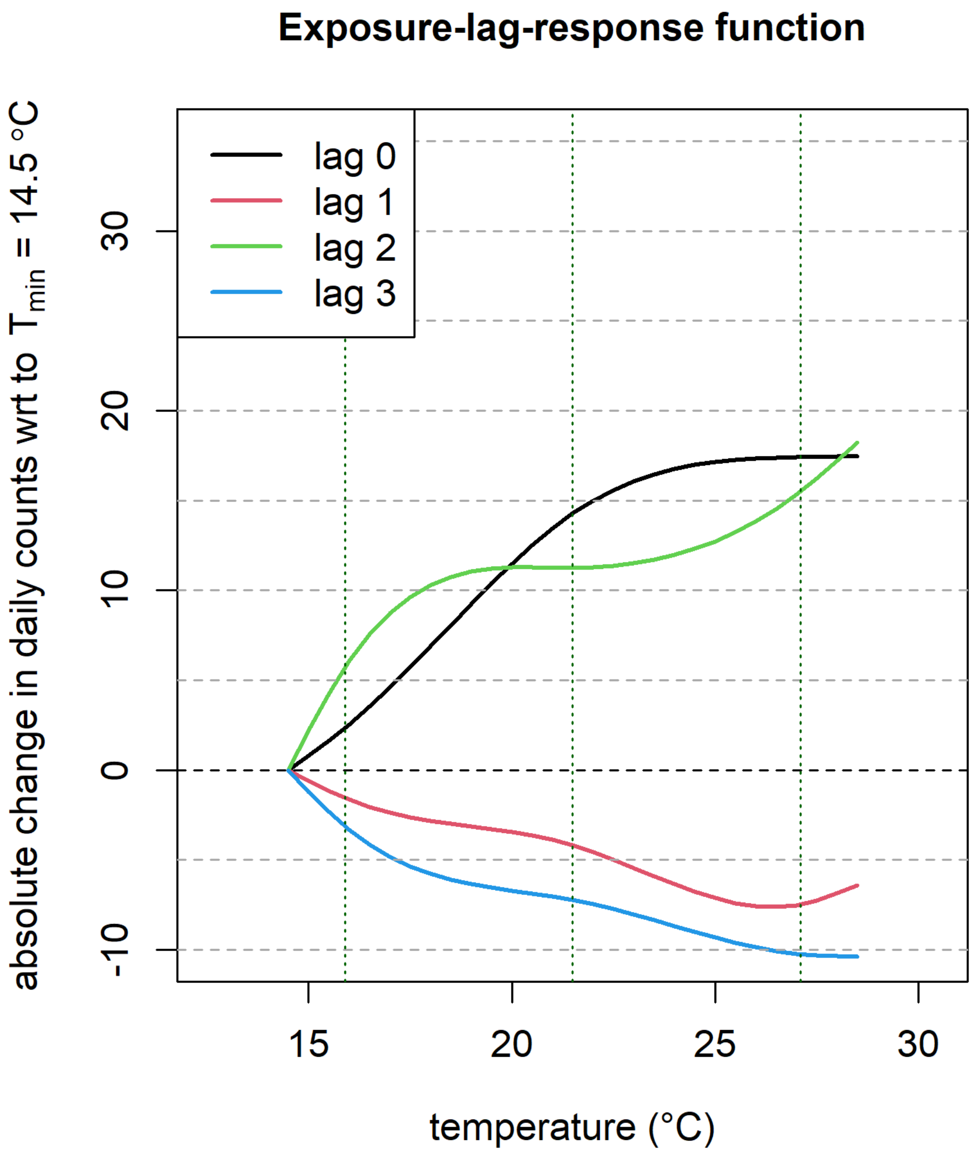


Exposure–lag–response functions describing the association between daily mean temperature and the absolute change in daily emergency department (ED) visit counts in Lugano.

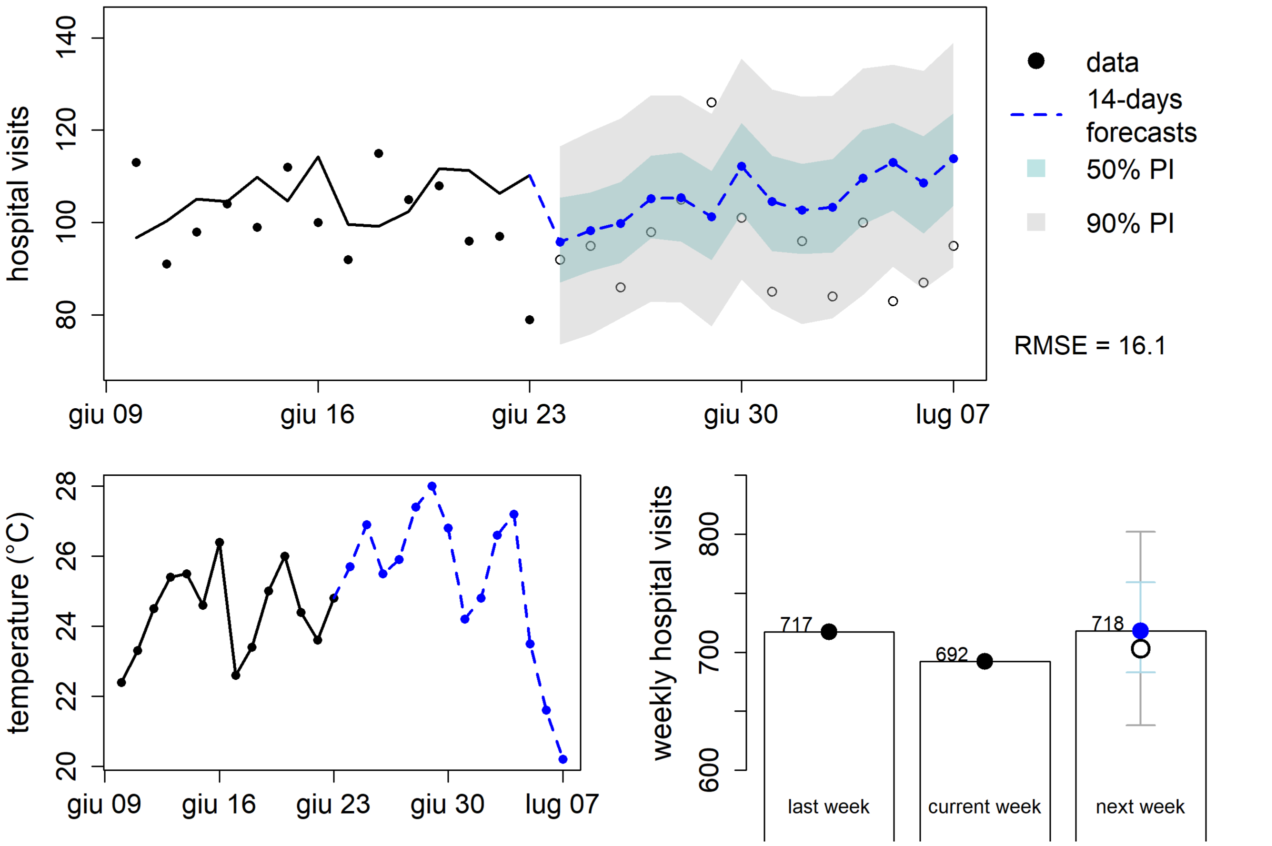


**Forecast of emergency department visits in Lugano during the June 2025 heatwave (HW1).**
The upper panel shows observed daily hospital visits from 9 to 23 June 2025 (black points and line) and 14-day forecasts from 24 June to 7 July 2025 (blue dashed line) with 50% and 90% prediction intervals. The forecast period includes the heatwave observed in Lugano from 24 to 30 June 2025. The lower left panel shows the corresponding daily mean temperature (°C). The lower right panel summarizes weekly hospital visits, comparing the previous week, the current week, and the forecast for the following week.

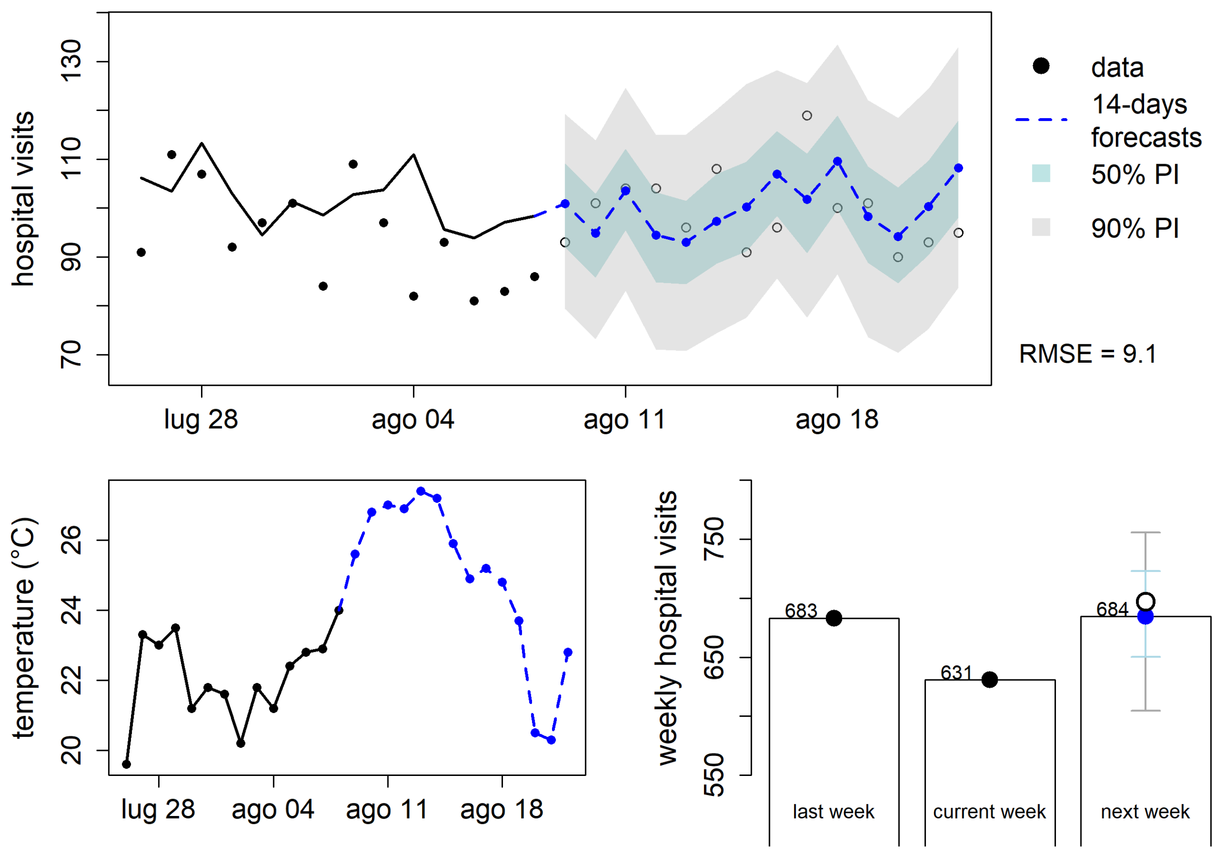


**Forecast of emergency department visits in Lugano during the August 2025 heatwave (HW2).** The upper panel shows observed daily hospital visits from 28 July to 8 August 2025 (black points and line) and 14-day forecasts from 9 to 22 August 2025 (blue dashed line) with 50% and 90% prediction intervals. The forecast period includes the heatwave observed in Lugano from 9 to 15 August 2025. The lower left panel shows the corresponding daily mean temperature (°C). The lower right panel summarizes weekly hospital visits, comparing the previous week, the current week, and the forecast for the following week.

<u>**This figure is already presented in the manuscript as Figure 3.**</u>

*Forecast errors (cross validation). Temperature threshold: 25°C*

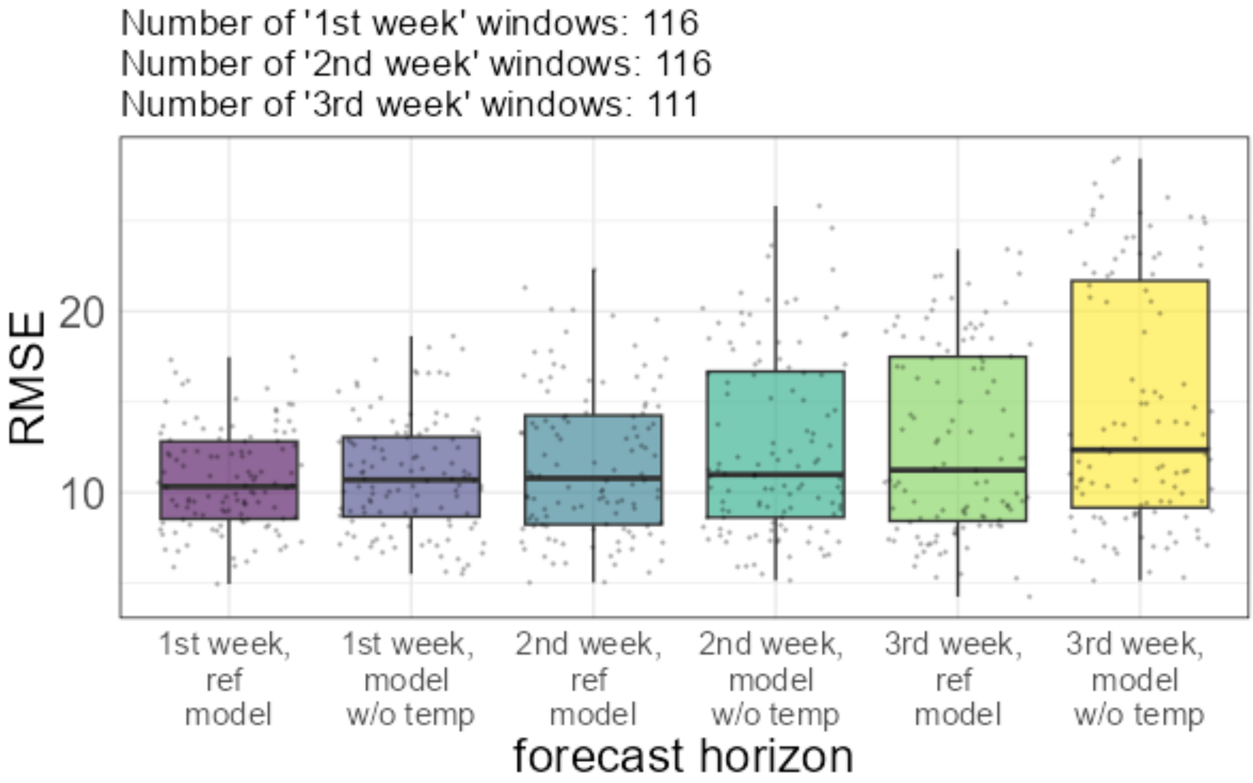

### MENDRISIO (STABIO)

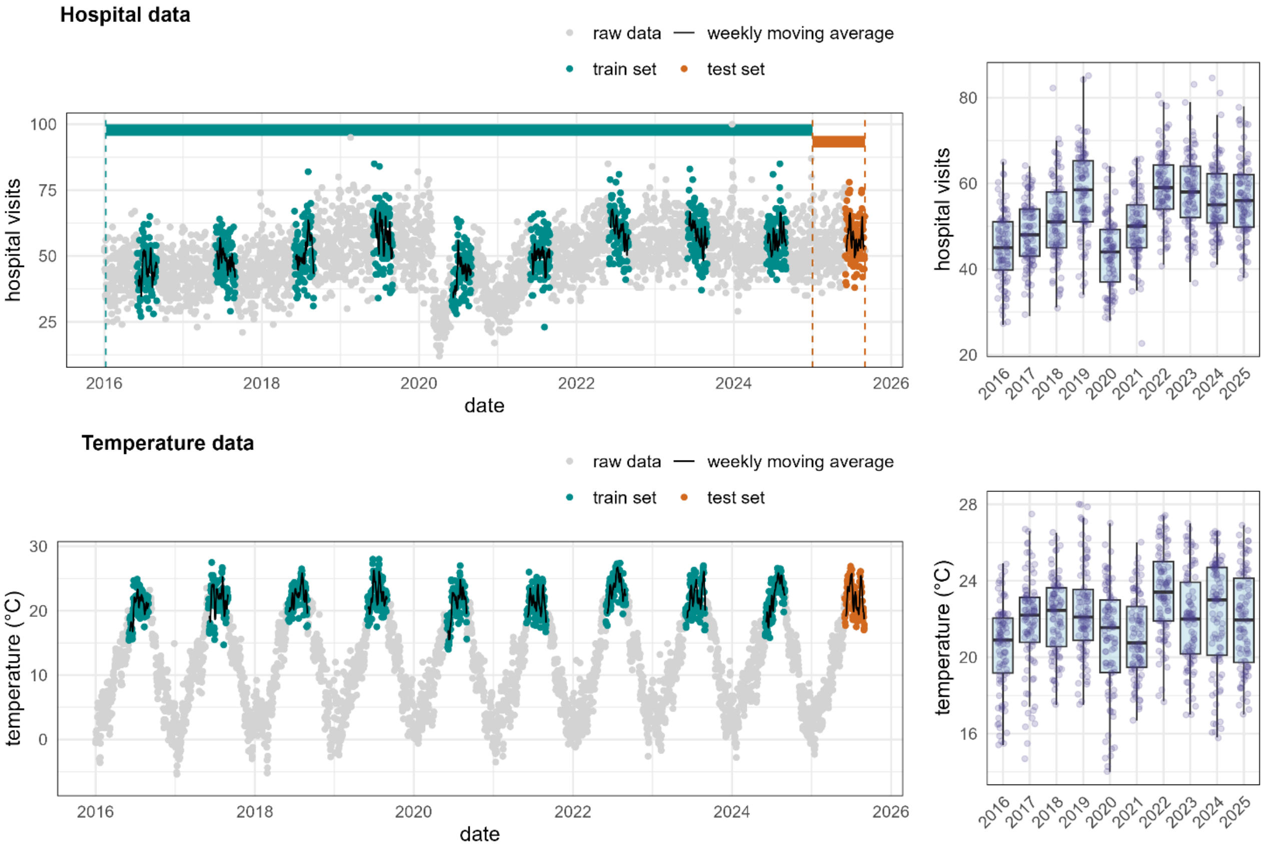


Daily mean temperature (°C) in Mendrisio (Stabio station) during the summer period (1 June–31 August) from 2016 to 2025. The dashed line represents the 25 °C threshold; red points indicate days when the temperature exceeded this value, identifying potential heatwave conditions.

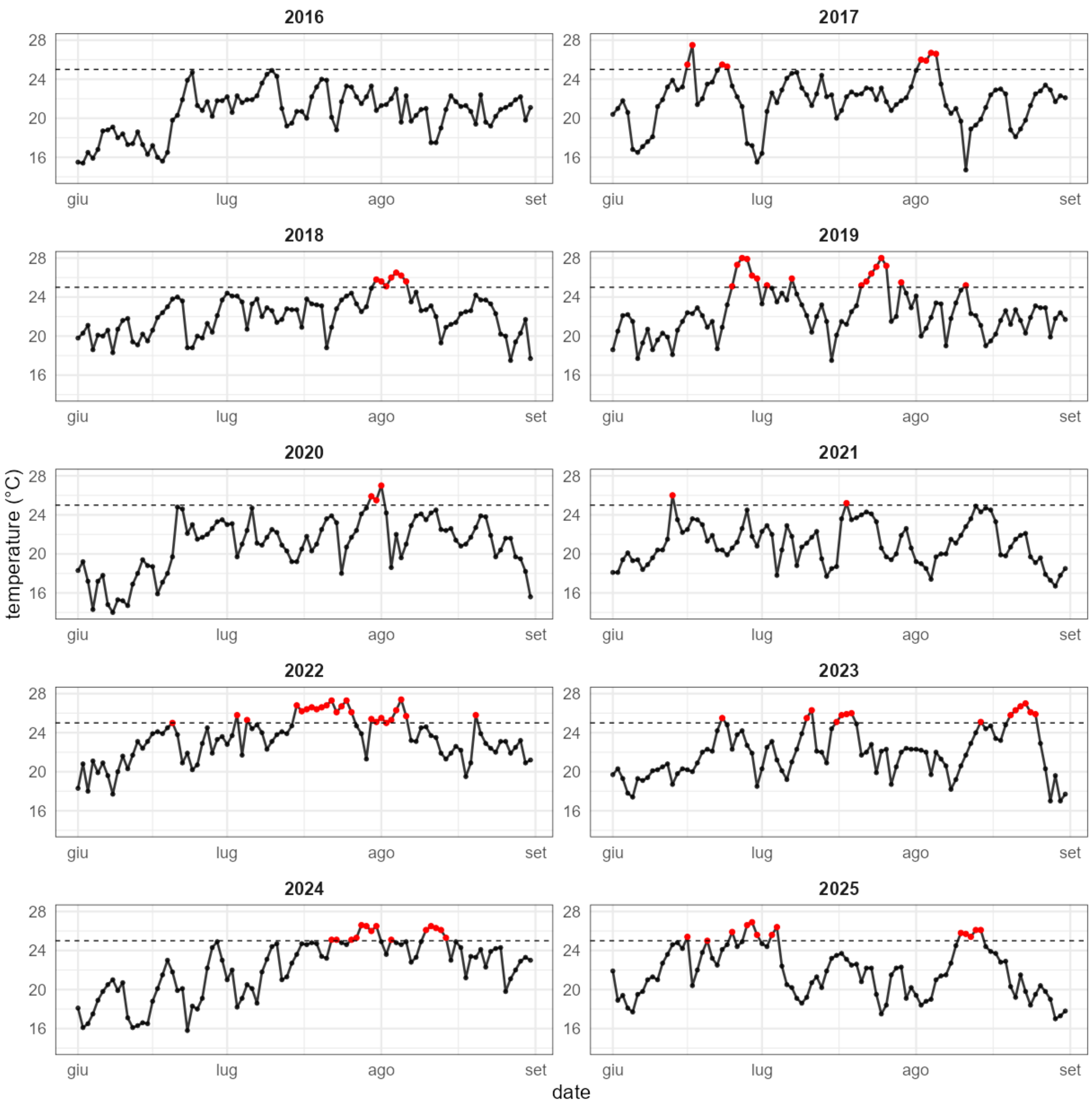


Daily mean temperature (°C) in Mendrisio (Stabio station) during the summer period (1 June–31 August) from 2016 to 2025. The dashed line represents the 25 °C threshold; red points indicate days when the temperature exceeded this value.

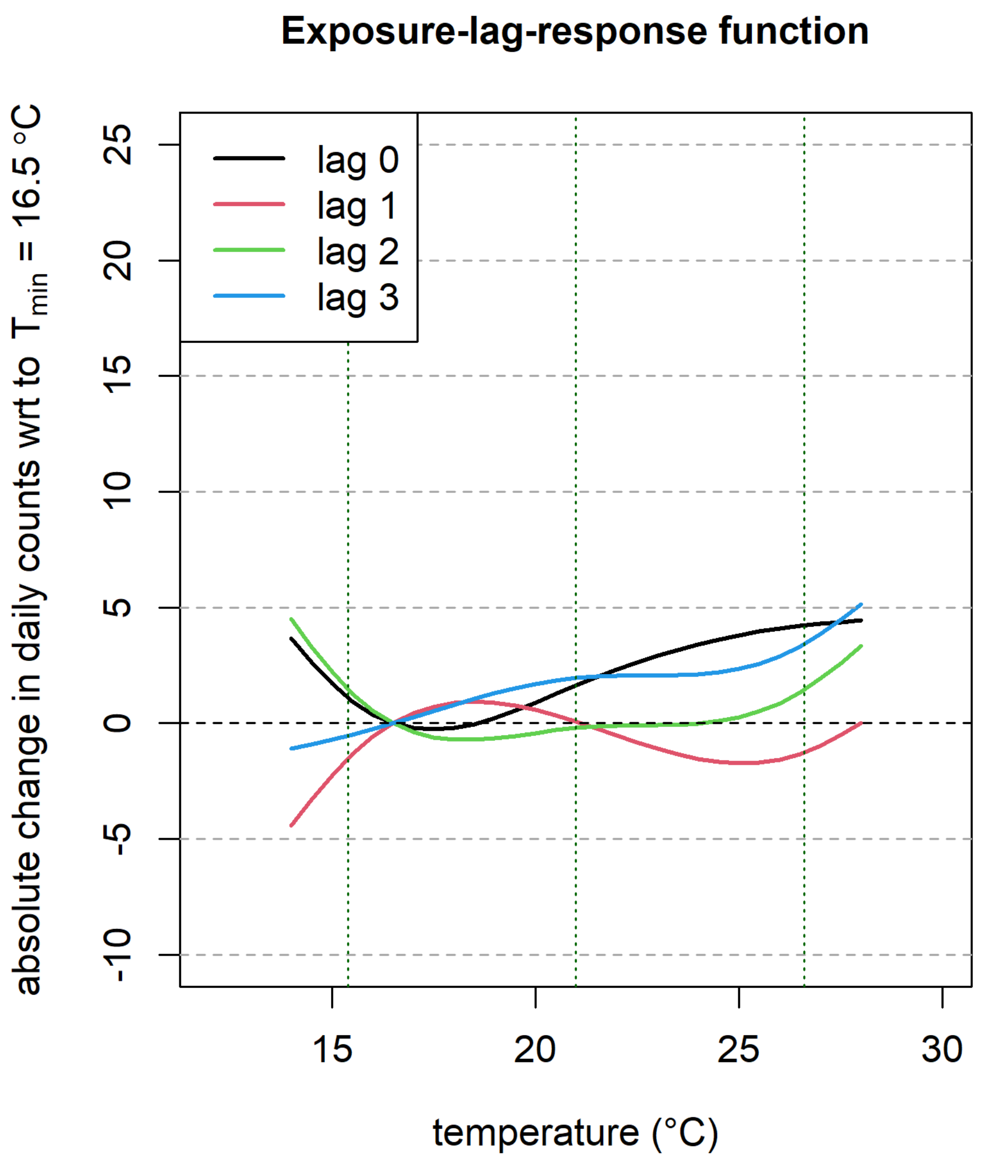


Exposure–lag–response functions describing the association between daily mean temperature and the absolute change in daily emergency department (ED) visit counts in Mendrisio.

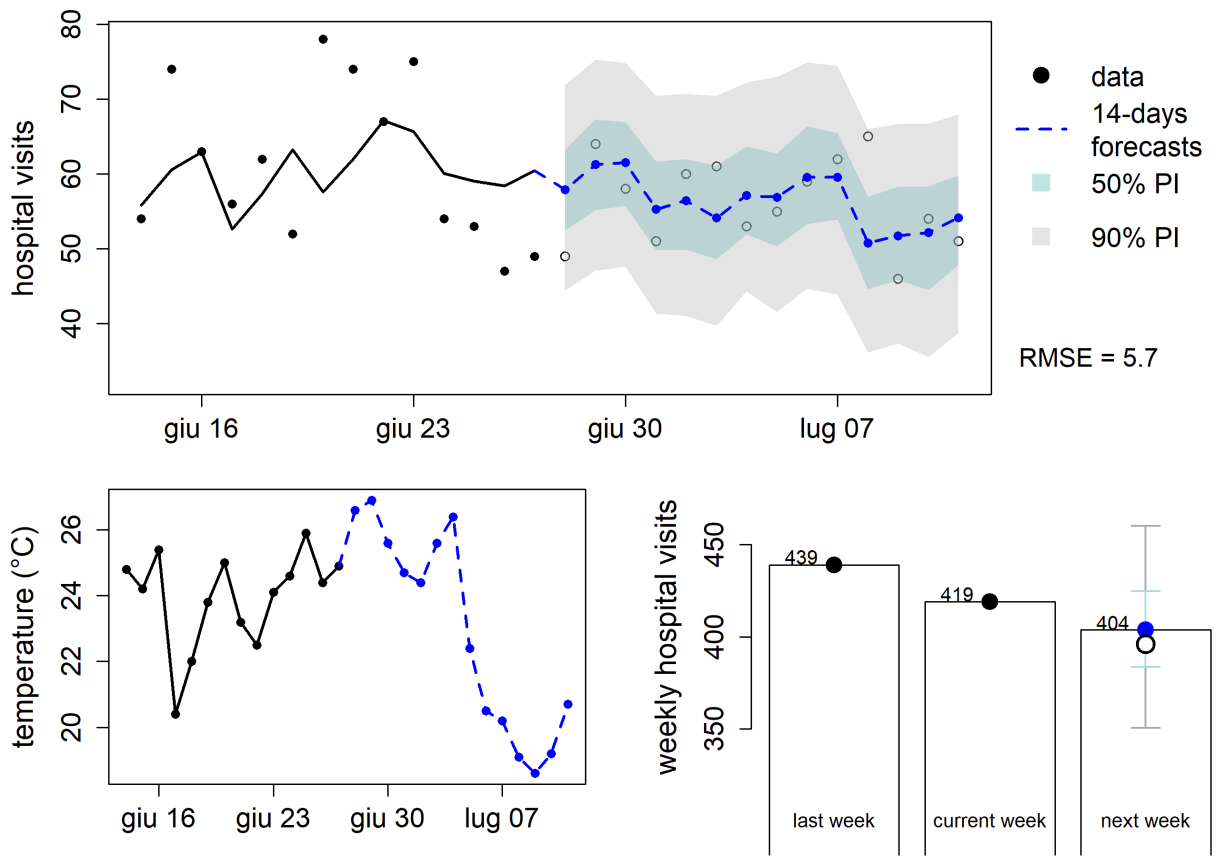


**Forecast of emergency department visits in Mendrisio during the June 2025 heatwave (HW1).** The upper panel shows observed daily hospital visits from 12 to 27 June 2025 (black points and line) and 14-day forecasts from 28 June to 11 July 2025 (blue dashed line) with 50% and 90% prediction intervals. The forecast period includes the heatwave observed in Mendrisio from 28 to 30 June 2025. The lower left panel shows the corresponding daily mean temperature (°C). The lower right panel summarizes weekly hospital visits, comparing the previous week, the current week, and the forecast for the following week.

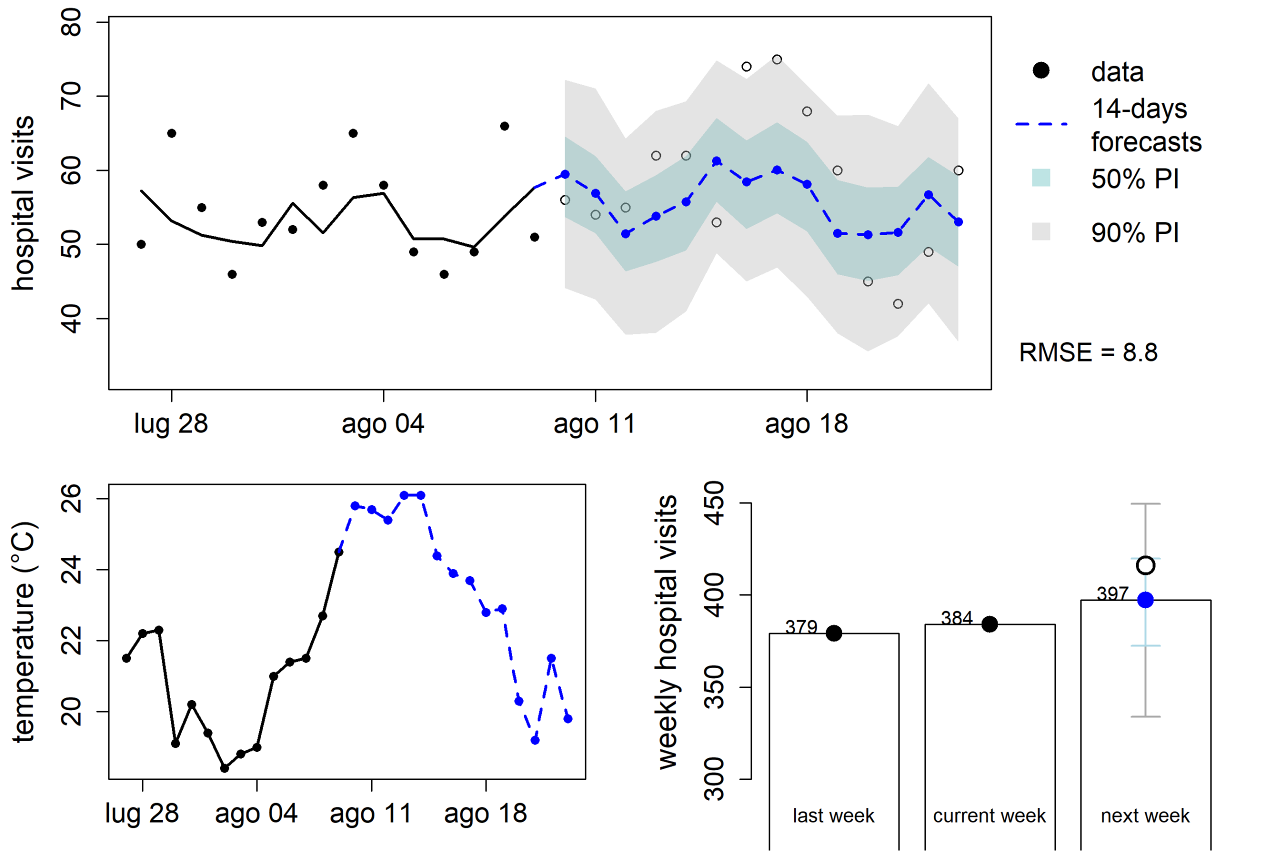


**Forecast of emergency department visits in Mendrisio during the August 2025 heatwave (HW2).** The upper panel shows observed daily hospital visits from 28 July to 9 August 2025 (black points and line) and 14-day forecasts from 10 to 23 August 2025 (blue dashed line) with 50% and 90% prediction intervals. The forecast period includes the heatwave observed in Mendrisio from 10 to 14 August 2025. The lower left panel shows the corresponding daily mean temperature (°C). The lower right panel summarizes weekly hospital visits, comparing the previous week, the current week, and the forecast for the following week

*Forecast errors (cross validation). Temperature threshold: 25°C*

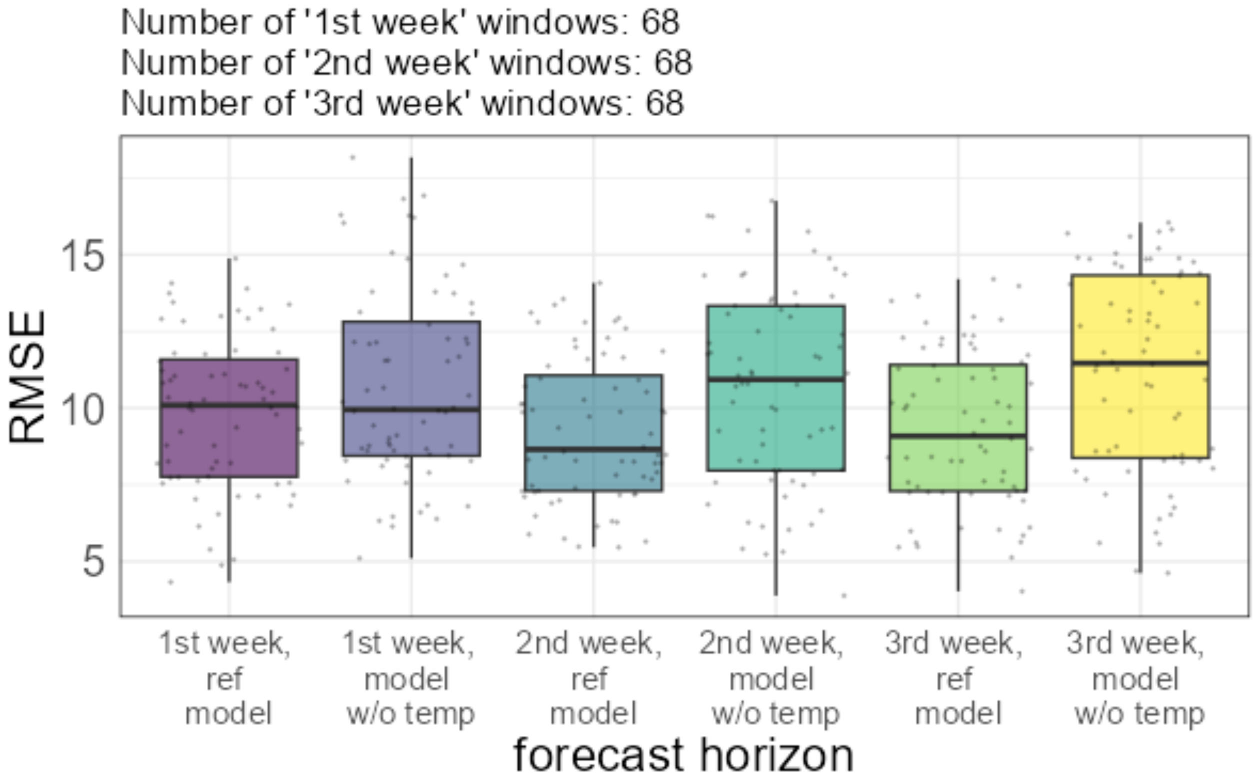
